# Understanding the dynamic relation between wastewater SARS-CoV-2 signal and clinical metrics throughout the pandemic

**DOI:** 10.1101/2022.07.06.22277318

**Authors:** Nada Hegazy, Aaron Cowan, Patrick M. D’Aoust, Élisabeth Mercier, Syeda Tasneem Towhid, Jian-Jun Jia, Shen Wan, Zhihao Zhang, Md Pervez Kabir, Wanting Fang, Tyson E. Graber, Alex E. MacKenzie, Stéphanie Guilherme, Robert Delatolla

## Abstract

Wastewater surveillance (WWS) of SARS-CoV-2 was proven to be a reliable and complementary tool for population-wide monitoring of COVID-19 disease incidence but was not as rigorously explored as an indicator for disease burden throughout the pandemic. Prior to global mass immunization campaigns and during the spread of the wildtype COVID-19 and the Alpha variant of concern (VOC), viral measurement of SARS-CoV-2 in wastewater was a leading indicator for both COVID-19 incidence and disease burden in communities. As the two-dose vaccination rates escalated during the spread of the Delta VOC in Jul. 2021 through Dec. 2021, relations weakened between wastewater signal and community COVID-19 disease incidence and maintained a strong relationship with clinical metrics indicative of disease burden (new hospital admissions, ICU admissions, and deaths). Further, with the onset of the vaccine-resistant Omicron BA.1 VOC in Dec. 2021 through Mar. 2022, wastewater again became a strong indicator of both disease incidence and burden during a period of limited natural immunization (no recent infection), vaccine escape, and waned vaccine effectiveness. Lastly, with the populations regaining enhanced natural and vaccination immunization shortly prior to the onset of the Omicron BA.2 VOC in mid-Mar 2022, wastewater is shown to be a strong indicator for both disease incidence and burden. Hospitalization-to-wastewater ratio is further shown to be a good indicator of VOC virulence when widespread clinical testing is limited. In the future, WWS is expected to show moderate indication of incidence and strong indication of disease burden in the community during future potential seasonal vaccination campaigns.

**Highlights:**

- Need to elucidate interpretation of CoV-2 WWS for seasonal vaccination campaigns.
- WWS to incidence relation weakens with peak natural and vaccination immunization.
- WWS to hospitalization remains strong with natural and vaccination immunization.
- WWS as indicator of hospitalization during future seasonal vaccination campaigns.
- WWS/hospitalization as indicator of VOC virulence with limited clinical testing.

## 1 Introduction

Wastewater surveillance (WWS) is a population-wide approach that uses municipal wastewater for real-time surveillance of public health status. Historically, WWS has played a role in the Global Polio Eradication Initiative (GPEI) in 1988 to monitor poliovirus outbreaks around the world (Asghar et al., 2014). Wastewater monitoring has further proven to effectively forecast disease outbreaks caused by norovirus (Prevost et al., 2015; Santiso-Bellón et al., 2020) and hepatitis A (Hellmér et al., 2014). During the spread of the wildtype and the B.1.1.7 (Alpha) Coronavirus disease 2019 (COVID-19) between March 2020 and May 2021, WWS rapidly emerged as a reliable and complementary indicator of severe acute respiratory syndrome coronavirus 2 (SARS-CoV-2) infections in various communities around the world.

At the time of writing this article (Jun. 2022), WWS of COVID-19 is active worldwide in nearly 60 countries at over 3000 sites (COVIDPoops19, 2022; Naughton et al., 2021) and is playing an increasingly significant role in the real-time monitoring of SARS-CoV-2 infection dynamics throughout the world (Ahmed et al., 2020; Bar-Or et al., 2020; Gonzalez et al., 2020; Haramoto et al., 2020; Kocamemi et al., 2020; La Rosa et al., 2021; Medema et al., 2020; Nemudryi et al., 2020; Peccia et al., 2020; Randazzo et al., 2020; Rimoldi et al., 2020; Sherchan et al., 2020; Wu et al., 2021; Wurtzer et al., 2020; Zhang et al., 2021). Various studies identified a significant correlation between the wildtype and the early variant of concern (VOC) B.1.1.7 (Alpha) (dominant in Canada from Mar. 2021 to Jul. 2021) SARS-CoV-2 RNA measurements in wastewater and reported COVID-19 clinical reporting of nasopharyngeal testing results with a 0 – 4 day lead time (time-lag between viral detection in wastewater and onset of symptoms), thereby establishing wastewater signal of SARS-CoV-2 RNA as an effective early indicator for disease incidence (D’Aoust et al., 2021a; Gerrity et al., 2021; Gonzalez et al., 2020; Graham et al., 2021; Medema et al., 2020; Peccia et al., 2020). Additionally, a strong relation between wastewater measurements of wildtype and the B.1.1.7 (Alpha) VOC SARS-CoV-2 RNA viral signal and hospitalization of patients with severe COVID-19 symptoms was reported with a lead time of 19 – 21 days (Peccia et al., 2020; Saguti et al., 2021) and 3 – 9 days based on mathematical modelling predictions (Galani et al., 2022; Kaplan et al., 2021). Mathematical modelling predictions also demonstrated wastewater is an indicator for intensive care unit (ICU) admissions with a lead time of 3 – 8 days (Galani et al., 2022). With such a considerable time gap between viral measurements in wastewater and reported hospitalizations and ICU admissions, the application of WWS for COVID-19 surveillance provided to be an early prediction of health care demands for COVID-19-related symptoms, while providing insight into disease burden through its relation to hospitalizations and ICU admissions.

With the progression of the COVID-19 pandemic and the predomination of new VOCs such as the B.1.617.2 (Delta) VOC (dominant in Canada from Jul. 2021 to Dec. 2021), B.1.1.529.1 (first Omicron BA.1 sub-lineage dominant in Canada from Dec. 2021 to Feb. 2022), and B.1.1.529.2 (second Omicron BA.2 sub-lineage dominant in Canada since Mar. 2022 from the time of writing this article (Jun. 2022)) along with the initiation of mass vaccination immunization campaigns in many countries, the relation between wastewater viral signal and COVID-19 positive cases was shown to change (D’Aoust et al., 2022; Xiao et al., 2021), thus affecting the predictive abilities of WWS for disease incidence in the community. In addition, the relation between wastewater signal and hospital admissions, ICU admissions, and COVID-19 caused deaths during the progression of the pandemic has not currently been as rigorously explored compared to positive cases, and hence the effects of the relation to disease burden throughout the pandemic is less evident. Prior to initiation of mass immunization campaigns across the world, 57% of symptomatic individuals were likely to exhibit moderate COVID-19 symptoms, and 0.9% were likely to display severe symptoms (Inokuchi et al., 2021). Following mass immunization campaigns and concurrently with the emergence of the more transmissible B.1.617.2 (Delta) VOC, and with over 60% of the total Canadian population fully vaccinated against COVID-19 (two approved COVID-19 doses) by the end of summer 2021, the development of severe symptoms that would require treatment in hospitals lowered (Levine-Tiefenbrun et al., 2021; Paredes et al., 2021). Further into the pandemic, over 16 weeks having passed since 60% of the total population in Canada received two doses of the COVID-19 vaccine (Aug. 3^rd^, 2021) (Mathieu et al., 2021), the increasingly infectious B.1.1.529.1 (Omicron BA.1) was detected as a new VOC (Nov. 26^th^, 2021) entering the community and rapidly overtook the B.1.617.2 (Delta) VOC (WHO, 2021). The effectiveness of the widely used mRNA vaccine against symptomatic disease from the B.1.1.529.1 (Omicron BA.1) VOC was found to wane over time from 45.9 to 88% after 2 – 9 weeks from receiving two doses of mRNA vaccine to only 14.9 – 36.3% after ≥ 25 weeks (Andrews et al., 2022; Iorio A. et al., 2022; Lin et al., 2021; Tartof et al., 2021). The mRNA-based vaccine booster (third dose) was found to once more increase vaccine-induced immunization against symptomatic disease up to 75% at 2 weeks, 56.6% at 4 to 5 weeks, and 43.7% at 10 to 11 weeks from third dose reception (Iorio A. et al., 2022). As such, in light of the B.1.1.529.1 (Omicron BA.1) VOC spread in Canada, vaccine booster dose rollout increased rapidly between Jan. and Feb. 2022. As of Feb. 27^nd^, 2022, 45.74% of the total population in Canada received a booster vaccine dose (Government of Canada, 2022); optimizing vaccination immunization in almost half of the population prior to entering the sixth surge of the COVID-19 pandemic by mid-March, 2022, driven by the B.1.1.529.2 (Omicron BA.2) VOC.

Presently, a gap of knowledge exists with respect to WWS functioning as a predictor and indicator of disease incidence and/or disease burden during periods of peak vaccination immunization (with a significant portion of the population receiving two vaccine doses 2 – 4 weeks prior to or during the onset of a predominating VOC), and during periods of limited natural immunization and waning vaccination immunization (with a significant portion of the population last receiving two vaccine doses beyond 20 weeks of the onset of the predominating VOC). With significant WWS efforts ongoing across the globe for over 24 months along with the emergence of new VOCs and initiation of nationwide vaccination immunization, in addition to vaccination and booster campaigns, there is a present opportunity to explore how WWS reflects clinical epidemiological metrics based on timely distribution of vaccines (limited waning of vaccine effectiveness), and whether the vaccine is effective for the dominant VOC (limited vaccine-escape). It was previously proposed by *Xiao et al. (2021)* to use wastewater signal/laboratory positive cases (WC) as a means for estimating true disease incidence and virulence (Xiao et al., 2021). However, continuous and population-wide representative SARS-CoV-2 diagnosis using polymerase chain reaction (PCR) testing has presently become less applied in many countries due to the economic impact of testing. Rapid antigen testing has become widely used globally for mass testing, with limited reporting of results. As such, there currently exists a high likelihood of underreporting of COVID-19 laboratory positive cases in many countries, where small cohorts of populations are eligible for PCR testing and hence laboratory positive testing of COVID-19 becoming a less reliable metric of community incidence. Therefore, the ability to continue to relate WWS measurements to population-wide, representative laboratory positive testing results and community incidence has diminished and will likely continue to diminish across the world. This highlights the need to review and study the previous two years of WWS measurements to clinical testing relations and to continue to monitor the relation between WWS measurements and disease burden via hospitalization. Further, there remains an opportunity to determine whether the relationships between wastewater and hospital admissions are a good indicator of VOC virulence in the absence of population-wide clinical testing. The objective of this research is to evaluate the relationship between SARS-CoV-2 wastewater viral measurements and four public health metrics throughout the pandemic in two long-standing monitored locations in Canada. The four specific public health metrics that will be related to WWS are i) laboratory positive cases, ii) new hospital admissions, iii) ICU admissions, and iv) COVID-19-caused deaths. These relations will be evaluated during the period preceding significant natural and vaccinated immunity and following significant natural and vaccinated immunity to elucidate the use of wastewater in the future for SARS-CoV-2 surveillance and the use during future potential seasonal vaccination campaigns.

## 2 Materials and methods

### 2.1 Wastewater sampling locations and characteristics

Twenty-four-hour composite samples of primary clarified sludge were collected from the City of Ottawa’s (Ontario, Canada) sole water resource recovery facilities (WRRF), Robert O. Pickard Environmental Centre (ROPEC), and from the City of Hamilton’s (Ontario, Canada) largest WRRF that services over 80% of the community’s population (Woodward Avenue Treatment Plant). Primary clarified sludge samples were first collected on April 8^th^, 2020, from the Ottawa WRRF, and July 18^th^, 2020, from the Hamilton WRRF. Daily sample collection and SARS-CoV-2 RNA extraction were ongoing in Ottawa from September 10^th^, 2020, up until the time of writing of this article (Jun. 2022). A sampling frequency of 3 to 4 days per week was performed in Hamilton. The Ottawa WRRF services approximately 936,382 people and has an average daily capacity of 545 million litres. The Hamilton Woodward WRRF services approximately 485,017 people and has an average daily capacity of 409 million litres. Composite samples from both WRRFs were collected, immediately kept on ice and transported to the University of Ottawa laboratory for analysis. Typical primary sludge characteristics from the Ottawa and Hamilton (Woodward) WRRFs across 2020 are shown in Table 1. It is noted that the characteristics of the primary sludges do not change significantly in the cities year to year.

**Table 1:**
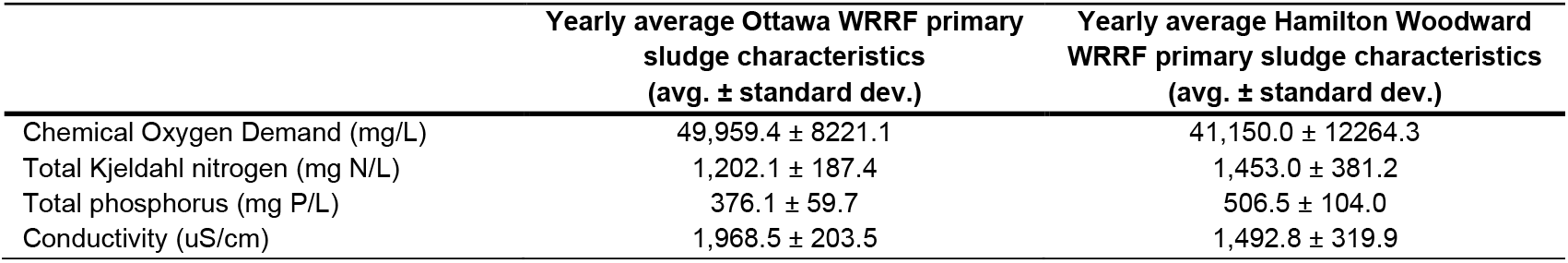
Yearly average primary sludge characteristics (2020) at Ottawa WRRF and Hamilton Woodward WRRF

### 2.2 RNA extraction and RT-qPCR quantification of SARS-CoV-2 in wastewater

40 mL of well-mixed, 24-hour composite primary sludge samples from Ottawa and Hamilton were concentrated by centrifugation at 10,000 x g for 45 minutes at 4°C. The resulting pellet was well-mixed and 0.250 ± 0.05 g of the resulting pellet was immediately processed for viral extraction using Qiagen’s RNeasy PowerMicrobiome extraction kit (PN 26000-50, MD, USA) on a QIAcube Connect automated extraction platform with a modified methodology previously described (D’Aoust et al., 2021b, 2021c). Quantification of SARS-CoV-2 RNA was performed by RT-qPCR targeting the N1 and N2 gene regions (D’Aoust et al., 2021b, 2021c). Additional RT-qPCR quantifications of the pepper mild mottle virus (PMMoV) were performed with 1/10 dilution of the same samples. Each PCR reaction consisted of 3 µL of RNA template. The primer and probe used in this study are shown in Supplemental Table S1. To check for inhibition, the samples were diluted by a factor of four and then a factor of ten and were compared with undiluted samples for the corresponding decline in PMMoV signal. The assay limit of detection (ALOD; ≥ 95% detection) is approximately 2 copies/reaction. RNA extraction and RT-qPCR were performed in separate laboratories in Class 2 biosafety cabinets to avoid contamination. The measured copies of SARS-CoV-2 (N1 and N2 gene regions) per reaction were normalized against measured PMMoV copies from respective samples.

### 2.3 Epidemiological data

Daily laboratory confirmed positive COVID-19 cases by report date and daily new hospital admissions due to COVID-19 infection in Ottawa between April 8^th^, 2020, and Feb 22^nd^, 2022, were obtained from Public Health Ontario’s COVID-19 online data tool (Public Health Ontario, 2022a) and Ottawa Public Health’s online COVID-19 dashboard (Ottawa Public Health, 2022), respectively. Laboratory positive COVID-19 cases and daily hospital admissions from COVID-19 infection in Hamilton between July 18^th^, 2020 and Feb 13^nd^, 2022, were obtained from the City of Hamilton online COVID-19 database (City of Hamilton, 2022). Further epidemiological daily measurements representing disease burden including ICU admissions and reported deaths due to COVID-19 complications were obtained from the Ottawa Public Health’s online COVID-19 dashboard (Ottawa Public Health, 2022) and from Hamilton’s public health dashboard (City of Hamilton, 2022). Vaccination information for both Ottawa and Hamilton was obtained from Public Health Ontario’s COVID-19 tool (Public Health Ontario, 2022a).

### 2.4 Statistical analysis

Spearman’s Rank correlation analyses between the 5-day and 3-day midpoint average of the wastewater viral signal measurements in Ottawa and Hamilton, respectively, and the 5-day midpoint average of the four sets of epidemiological data (laboratory positive cases, hospital admissions, ICU admissions, and deaths). Pearson’s R values ranging from 0 – 0.19 would suggest very weak correlations, 0.2 – 0.39 suggest weak correlations, 0.4 – 0.59 suggest a moderate correlation, 0.6 – 0.79 suggest strong correlation and 0.8 – 1 suggest an extraordinarily strong correlation. To determine whether a lag time (Δt), measured in days, exists between the wastewater SARS-CoV-2 measurements and epidemiological metrics, a time step analysis was performed by offsetting wastewater SARS-CoV-2 measurements forward in time by a period of 1 – 25 days. The forward time step (lag time of the epidemiological metric) with the strongest correlation between the data sets, along with visual alignment in recorded peaks, was considered when selecting the optimal lag time between wastewater measurements and epidemiological data.

## 3 Results and discussion

### 3.1 Relation between WWS signal and epidemiological metrics prior to significant natural and vaccination immunization, wildtype and Alpha surges

From Apr. 8^th^, 2020 to Aug. 29^th^, 2021, in Ottawa, and July 14^th^, 2020 to Oct. 9^th^, 2021, in Hamilton, less than 70% of the total population in both cities qualified as “fully vaccinated” (two doses of approved vaccine) against COVID-19 (Public Health Ontario, 2022a). Prior to significant community vaccination immunization, the asymptomatic proportion is believed to have ranged from 17.9% to 57% relative to symptomatic individuals (Kimball et al., 2020; Mizumoto et al., 2020; Sah et al., 2021). The proportion of symptomatic individuals likely to display moderate and severe illness are 56.7% and 0.9%, respectively (Inokuchi et al., 2021). A time step analysis demonstrates a good relationship between the measured SARS-CoV-2 wastewater viral signal (normalized with PMMoV) and laboratory positive cases and hence community disease incidence, with a 5-day lead time in Ottawa (Δt = 5d) (R = 0.821) and a 10-day lead time (Δt = 10d) in Hamilton (R = 0.655) when under 70% of both populations were fully immunized (Table 2, Figure 1 A and B). These findings are in agreement with reported observations from previous WWS work performed prior to mass vaccination immunization (D’Aoust et al., 2021a; Gerrity et al., 2021; Gonzalez et al., 2020; Graham et al., 2021; Medema et al., 2020; Peccia et al., 2020).

**Table 2:** Correlations (Pearson’s R) between normalized SARS-CoV-2 RNA signal (N1-N2 copies/copies PMMoV) and laboratory positive cases, hospital admissions, ICU admissions, and deaths in Ottawa and Hamilton, prior to significant natural and vaccination immunization.

| Ottawa |  | 5-day midpoint avg.<br>laboratory positive<br>cases | 5-day midpoint avg.<br>hospital admissions | 5-day midpoint avg.<br>ICU admissions | 5-day midpoint avg.<br>deaths |
| --- | --- | --- | --- | --- | --- |
| | Lead time ($\Delta t$) | 5d | 10d | 10d | 19d |
|  | Pearson's R | 0.821 | 0.808 | 0.708 | 0.730 |
| Hamilton |  | 3-day midpoint avg.<br>laboratory positive<br>cases | 3-day midpoint avg.<br>hospital admissions | 3-day midpoint avg.<br>ICU admissions | 3-day midpoint avg.<br>deaths |
| | Lead time ($\Delta t$) | 10d | 14d | 14d | 20d |
|  | Pearson's R | 0.655 | 0.682 | 0.352 | 0.574 |

**Figure 1:**
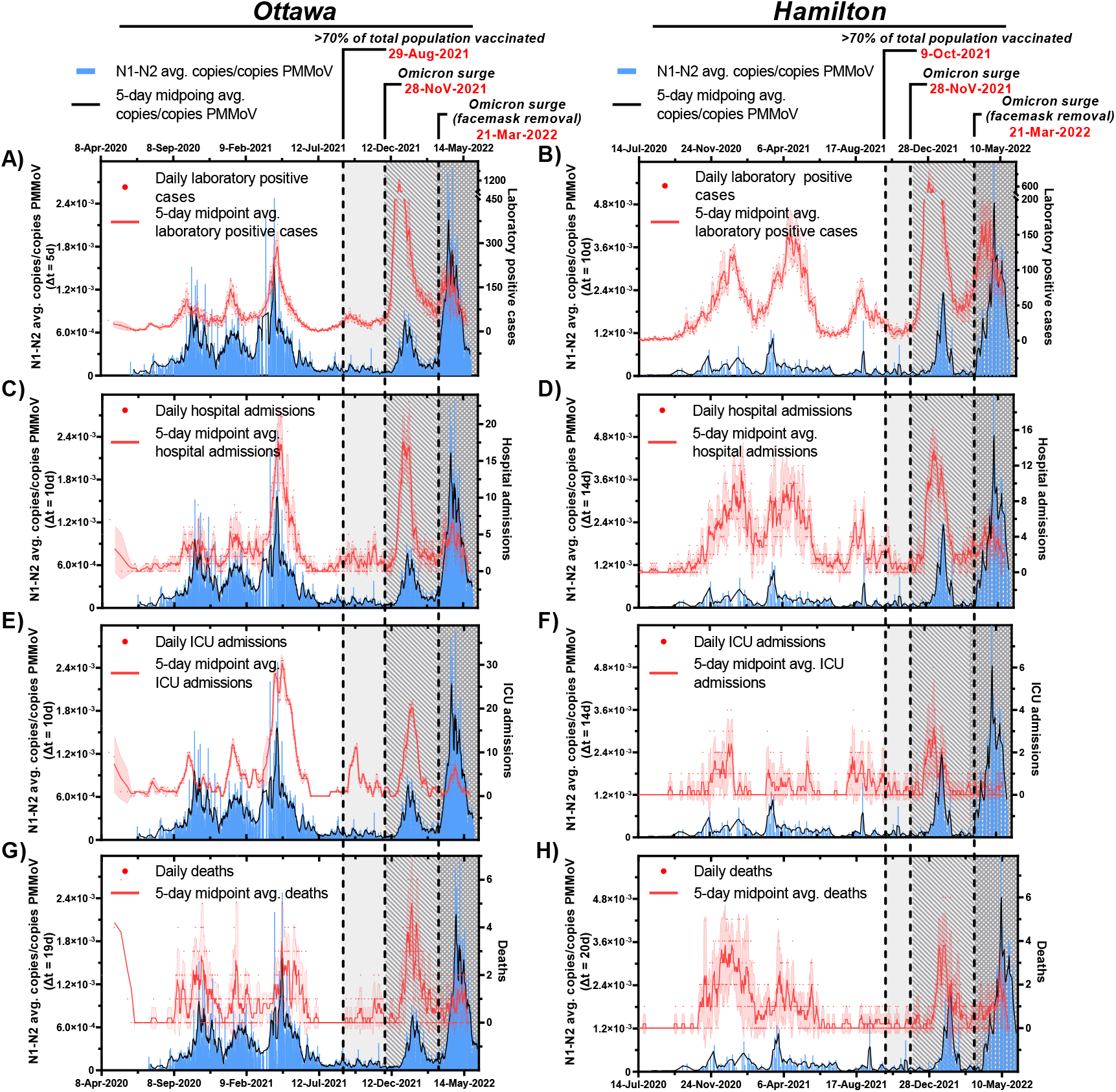
Relation between SARS-CoV-2 wastewater signal (advanced by respective “Δt” time lag on the x-axis) and A) clinical COVID-19 positive cases in Ottawa and B) clinical COVID-19 positive cases in Hamilton, C) hospital admissions in Ottawa and D) hospitalization admissions in Hamilton, E) ICU admissions in Ottawa and F) ICU admissions in Hamilton, and G) COVID-19 caused deaths in Ottawa, and H) COVID-19 caused deaths in Hamilton from Apr. 8^th^, 2020 to May. 26^th^, 2022. *\*Laboratory positive cases in Ottawa and Hamilton are underreported due to updated PCR eligibility in Ontario as of December 31*^*st*^, *2021*

A similar time step analysis further identified WWS as a good indicator for disease burden prior to significant vaccination immunization based on hospital admissions from COVID-19, and additionally based on ICU admissions, and deaths due to complications from COVID-19. A strong positive correlation exists between PMMoV normalized SARS-CoV-2 viral signal and hospital admissions with a 10-day lead time in the viral signal measurements in Ottawa (R = 0.808), and with a 14-day lead time in Hamilton (R = 0.682) (Table 2). This is further confirmed visually (Figure 1 C and D) and aligns with an observation from previous work demonstrating SARS-CoV-2 viral signal measurements preceding hospital admissions by over 10 days (D’Aoust et al., 2021a). PMMoV-normalized SARS-CoV-2 viral signal was further found to have a strong positive correlation with COVID-19 patients in ICU with Δt = 10d in Ottawa (R = 0.708); and a weak positive correlation exists between wastewater measurements and ICU admissions with Δt = 14d in Hamilton (R = 0.352) (Table 2, Figure 1 E and F, respectively). Finally, wastewater measurements maintain a strong positive relation with COVID-19-caused deaths in Ottawa with Δt = 19d (R = 0.730) (Table 2, Figure 1 G) and a moderate positive correlation with reported deaths in Hamilton with Δt = 20d (R = 0.574) (Table 2). The weak correlations between wastewater signal and epidemiological metrics in the city of Hamilton compared to the city of Ottawa are partially attributable to a lower frequency of wastewater testing in the city of Hamilton. Although weak positive correlations were found between wastewater measurements and reported ICU admissions and deaths in Hamilton, an apparent similarity in the trends between wastewater measurements and reported deaths in Hamilton is visually confirmed (Figure 1 F and H). With these findings, it can be concluded that WWS provides affirmative information attributed to both incidence of COVID-19 infections, and disease burden prior to significant vaccination immunization.

### 3.2 Relation between WWS signal and epidemiological metrics post significant natural and vaccination immunization and prior to vaccination waning, Delta surge and prior to Omicron (B.1.1.529) surge

As of Aug. 29^th^, 2021, in Ottawa, and Oct. 9^th^, 2021, in Hamilton, ≥ 70% of the total population in these communities were characterized as having reached significant vaccination immunity (2 approved vaccine doses) against COVID-19 (Public Health Ontario, 2022a). By the time significant vaccination immunity was achieved, the B.1.617.2 (Delta) VOC has already replaced B.1.1.7 (Alpha) as the dominant VOC in most locations in Ontario. Compared to the preceding B.1.1.7 (Alpha) VOC, the B.1.617.2 (Delta) VOC is distinguished as more transmissible and virulent (Kannan et al., 2021; Y. Liu et al., 2022; Planas et al., 2021) with higher risk of hospitalization to unvaccinated individuals (Paredes et al., 2021). To establish whether significant vaccination immunization affects the predictive nature and relation of wastewater measurements to community incidence and disease burden, the relationships between wastewater and epidemiological metrics (laboratory positive cases, new hospital admissions, ICU admissions, and deaths) were further evaluated using a time step analysis once significant vaccination immunization to just before the initial detection of the B.1.1.529.1 (Omicron BA.1) VOC in Ontario in Nov. 28^th^, 2021 (Public Health Ontario, 2022b). With significant vaccination immunization just being achieved in the communities, the majority of SARS-CoV-2 infections are less likely to exhibit severe COVID-19 symptoms leading to hospitalization or fatality due to lower viral loads (Levine-Tiefenbrun et al., 2021). Furthermore, with vaccine efficacy against the B.1.617.2 (Delta) VOC being over 75% at 17 weeks since reception of a second COVID-19 mRNA vaccine dose (Iorio A. et al., 2022), waning immunity was assumed to not be prevalent in either Ottawa or Hamilton between Aug. 29^th^, 2021 (Ottawa)/Oct. 9^th^, 2021 (Hamilton) and prior to the onset of the Omicron BA.1 (B.1.1.529.1) VOC on Nov. 28^th^, 2021.

During the periods of high natural and vaccination immunization between Aug. 29^th^, 2021 (Ottawa)/Oct. 9^th^, 2021 (Hamilton) and Nov. 28^th^, 2021, and prior to vaccination waning, the wastewater viral signal along with epidemiological data fluctuated at low levels as the B.1.617.2 (Delta) VOC spread through in the cities (Figure 2). Weak to moderate correlations were observed between the PMMoV-normalized SARS-CoV-2 viral signal measurements in wastewater and laboratory positive cases (Table 3). The weak correlations showed time steps of Δt = 0d in Ottawa (R = 0.189) and Δt = 0d in Hamilton (R = -0.029). This is further visually confirmed with wastewater signals remaining at low levels despite slight increases in laboratory positive case counts in both Ottawa and Hamilton communities (Figure 2 A and B, respectively). This indicates that wastewater is no longer leading clinical COVID-19 positive case counts in a community with recent, significant vaccination immunity.

**Table 3:** Correlations between normalized SARS-CoV-2 RNA signal (N1-N2 avg. copies/copies PMMoV) and laboratory positive cases, hospital admissions, ICU admissions, and deaths in Ottawa and Hamilton, post significant natural and vaccination immunization during the onset of the B.1.617.2 (Delta) VOC.

| Over 70% of the total population was fully vaccinated, Aug. 30<sup>th</sup> (Ottawa)/Oct.9<sup>th</sup> (Hamilton), 2021 – Nov. 27<sup>th</sup>, 2021, Delta surge |  |  |  |  |  |
| --- | --- | --- | --- | --- | --- |
| Ottawa |  | 5-day midpoint avg. laboratory positive cases | 5-day midpoint avg. hospital admissions | 5-day midpoint avg. ICU admissions | 5-day midpoint avg. deaths |
| | Lead time ($\Delta t$) | 0d | 11d | 14d | 25d |
|  | Pearson's R | 0.189 | 0.364 | 0.316 | 0.501 |

| Hamilton |  | 3-day midpoint avg. laboratory positive cases | 3-day midpoint avg. hospital admissions | 3-day midpoint avg. ICU admissions | 3-day midpoint avg. deaths |
| --- | --- | --- | --- | --- | --- |
| | Lead time ($\Delta t$) | 0d | 5d | 7d | 23d |
|  | Pearson's R | -0.029 | 0.312 | 0.524 | 0.097 |

**Figure 2:**
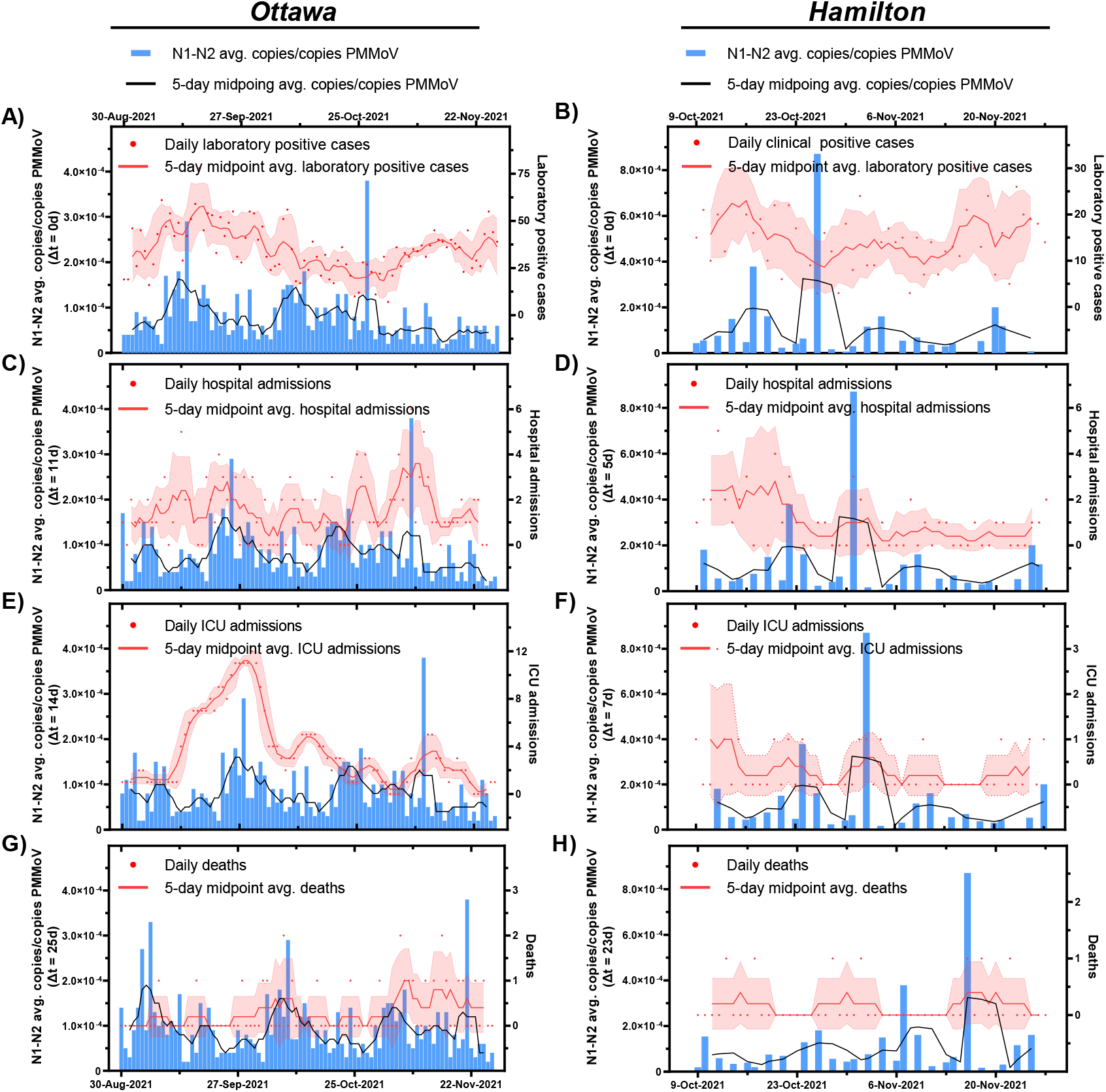
Relation between SARS-CoV-2 wastewater signal (advanced by respective “Δt” time lag on the x-axis) and A) clinical COVID-19 positive cases in Ottawa and B) clinical COVID-19 positive cases in Hamilton, C) hospital admissions in Ottawa and D) hospitalization admissions in Hamilton, E) ICU admissions in Ottawa and F) ICU admissions in Hamilton, and G) COVID-19 caused deaths in Ottawa, and H) COVID-19 caused deaths in Hamilton post 70% vaccination of total population and prior to Omicron surge (Aug. 30^th^, 2021 – Nov. 27^th^, 2021, in Ottawa, and Oct 9^th^, 2021 – Nov. 27^th^, 2021, in Hamilton)

While there was no change in PCR testing eligibility in Ontario during the B.1.617.2 (Delta) VOC surge, it is hypothesized that this loss in relation between wastewater signal and laboratory confirmed positive cases in both communities is attributed to reduced disease severity during the B.1.617.2 (Delta) VOC surge. As such, a similar time-step analysis was conducted between wastewater and epidemiological metrics reflective of disease burden (hospital admissions, ICU admissions, and deaths). A higher, but weak positive correlation exists between wastewater viral signals and hospital admissions at Δt = 11d in Ottawa (R = 0.364) (Table 3), and at Δt = 5d in Hamilton (R = 0.312) (Table 3). While the weak correlations between wastewater and the two clinical metrics are likely attributed to wastewater data fluctuating at low levels, the trends between the PMMoV-normalized viral signals and new hospital admission in Ottawa and Hamilton display better visual alignment at Δt = 11d (R = 0.364) (Figure 2 C) and Δt = 5d (R = 0.312) (Figure 2 D), respectively, compared to the overlayed trends of wastewater and laboratory positive cases (Figure 2 A and B). Furthermore, the weaker correlation between wastewater signal and hospital admissions in Hamilton, compared to the wastewater signals in Ottawa, may be due to the lower frequency of testing in Hamilton compared to Ottawa. A weak positive correlation was observed between PMMoV-normalized SARS-CoV-2 viral signal and ICU admissions in Ottawa at Δt = 14d (R = 0.316) (Table 3, Figure 2 E) and in Hamilton at Δt = 7d (R = 0.524) (Table 3, Figure 2 F). Finally, a moderate positive correlation was observed between PMMoV-normalized SARS-CoV-2 viral signal and reported daily deaths at Δt = 25d in Ottawa (R = 0.501) (Table 3, Figure 2 G), and at Δt = 23d in Hamilton (R = 0.097) (Table 3, Figure 2 H). Since no more than two COVID-19-related deaths were reported in Ottawa and Hamilton, the correlations between the wastewater measurements and reported deaths prior to the Omicron BA.1 (B.1.1.529.1) surge are presumed inconclusive. The results show a decline in the relation between wastewater and laboratory positive cases with ≥ 70% of the total population of both communities being recently fully vaccinated against COVID-19 and vaccination occurring prior to anticipated vaccine waning. Comparatively, hospitalization and ICU admissions exhibit a better relationship with the PMMoV-normalized viral signal measurements. These findings suggest the possibility of wastewater being a less strong lead indicator of disease incidence during possible, future COVID-19 seasonal vaccination campaigns while being a strong lead indicator of community disease burden.

### 3.3 Relation between WWS signal and epidemiological metrics during community vaccination waning, Omicron BA.1 (B.1.1.529.1) surge

The Omicron subvariant BA.1 (B.1.1.529.1) VOC was first detected in the Canadian province of Ontario on November 28^th^, 2021 (Public Health Ontario, 2022b). As of December 24^th^, 2021, 50.1% of laboratory positive cases were reportedly infected with Omicron BA.1 (B.1.1.529.1) in Ottawa (Figure S1 in Supplemental Material) (Ottawa Public Health, 2022) and as of December 26^th^, 2021, 50% of laboratory positive cases were reportedly infected with Omicron BA.1 (B.1.1.529.1) in Hamilton (Public Health Ontario, 2022a). One of the unique properties of the Omicron BA.1 (B.1.1.529.1) VOC is its higher vaccine-escape capabilities, resulting in reduced protection by vaccination immunization against onset disease severity and higher contagion compared to the Delta (B.1.617.2) VOC (Hogan et al., 2021; L. Liu et al., 2022; Smith-Jeffcoat et al., 2022; Wang et al., 2022). Additionally, at the onset of Omicron BA.1 (B.1.1.529.1), over 20 weeks have passed since ≥ 70% of the total population in the Ottawa and Hamilton communities achieved full vaccination immunization (two doses of approved vaccine). Evidence suggests waning of vaccine effectiveness against symptomatic disease from the Omicron BA.1 (B.1.1.529.1) from 75% at 2 weeks, to 14.9 – 36.3% at ≥ 25 weeks from the reception of a second COVID-19 mRNA vaccine dose (Andrews et al., 2022; Iorio A. et al., 2022; Lin et al., 2021; Tartof et al., 2021). Thus, subjecting both communities to waned vaccine-induced immunity against the highly infections Omicron BA.1 (B.1.1.529.1) VOC. This particularly puts the most frail groups in the community (the elderly, immunocompromised, and unvaccinated) at a greater risk of severe COVID-19-related complications and morbidity during the onset of Omicron BA.1 (B.1.1.529.1). A booster (third dose) of the mRNA-based vaccine was found to increase vaccine-induced immunization against symptomatic disease up to 75% at 2 weeks, 56.6% at 4 – 5 weeks, and 43.7 at 10 – 11 weeks from reception of the third vaccine dose (Iorio A. et al., 2022). As such, a booster COVID-19 vaccine dose campaign was initiated on Dec. 13, 2021, to provide improved vaccine-induced immunity against the Omicron BA.1 (B.1.1.529.1) VOC, with priority given to the most frail groups in the community. The Omicron BA.1 (B.1.1.529.1) VOC is further distinguished from the preceding VOCs by its shorter incubation period (the duration between infection and onset of symptoms) (Lee et al., 2021; Public Health Ontario, 2022c; Smith-Jeffcoat et al., 2022), and its shorter serial interval (the time from symptom onset in primary infector and symptom onset in secondarily infected case) (Backer et al., 2022; Public Health Ontario, 2022c). The implications of these novel characteristics on the relation between wastewater viral signal and the clinical metrics representing disease incidence and disease burden require further exploration.

With the onset of the Omicron BA.1 (B.1.1.529.1) VOC from November 28^th^, 2021, through February 22^nd^, 2022, measurements of the PMMoV-normalized SARS-CoV-2 RNA in wastewater from both Ottawa and Hamilton exhibited an abrupt rise along with laboratory positive case counts, new hospital admissions, ICU admissions, and deaths (Figure 1). At the peak of the Omicron BA.1 (B.1.1.529.1) surge, clinical case counts and hospital admissions reached a record high of 1176 laboratory positive cases and 23 hospital admissions after the wildtype and Alpha (B.1.1.7) surges (April 8^th^, 2020 – August 29^th^, 2021) (Figure 1). On December 31^st^, 2021, PCR testing eligibility in the Canadian province of Ontario changed from population-wide testing to only symptomatic and high-risk individuals being eligible for testing (Government of Ontario, 2021). Prior to this change in testing eligibility, a strong positive correlation between PMMoV-normalized SARS-CoV-2 viral signal and laboratory positive cases were observed in Ottawa at Δt = 0d (R = 0.861), and at Δt = 0d in Hamilton (R = 0.727) (Table 4). Following the December 31^st^, 2021, change in testing eligibility, the relation between wastewater viral signal and clinical reported cases has notably diminished due to reduced clinical testing and underreporting of the laboratory positive case counts in both Ottawa and Hamilton; this is further visually confirmed (Figure 3 A and B). On the contrary, PMMoV-normalized SARS-CoV-2 viral signal remained in strong agreement with new hospital admissions at Δt = 4d in Ottawa (R = 0.947), and at Δt = 3d in Hamilton (R = 0.837) (Table 4) from November 28^th^, 2021, through February 22^nd^, 2022; this is further visually confirmed (Figure 3 C and D), demonstrating WWS efforts consistently leading new hospital admissions. However, the lead time between PMMoV-normalized viral signal and new hospital admissions is notably shorter during the onset of the Omicron BA.1 (B.1.1.529.1) VOC in Ottawa and Hamilton compared to the onset of the Alpha (B.1.1.7) VOC that displayed lead times of 10 and 14 days in Ottawa and Hamilton, respectively (Table 2, Figure 1 A and B). This is likely attributed to the shorter incubation period ranging between 2 – 8 days in individuals infected with the Omicron BA.1 (B.1.1.529.1) VOC compared to the preceding VOCs (Lee et al., 2021; Smith-Jeffcoat et al., 2022). Furthermore, PMMoV-normalized SARS-CoV-2 viral signal measurements further maintains a strong positive relation with reported ICU admissions at Δt = 14d in Ottawa (R = 0.911), and at Δt = 2d in Hamilton (R = 0.673) (Table 4, Figure 3 E and F, respectively). The differences in the lead times of the PMMoV-normalized SARS-CoV-2 viral signals to the ICU admissions between Ottawa and Hamilton are likely attributed to the lower ICU admissions in Hamilton compared to Ottawa (Figure 3 E and F, respectively). Finally, a strong positive correlation further exists between PMMoV-normalized SARS-CoV-2 viral signal and reported deaths at Δt = 18d in Ottawa (R = 0.905), and at Δt = 9d in Hamilton (R = 0.663) (Table 4, Figure 3 G and H), supporting the indication of disease burden. Thus, findings suggest that during the onset of the Omicron BA.1 (B.1.1.529.1) VOC with its vaccine-escape capabilities, wastewater viral signal measurements were strong indicators of disease burden in both the Ottawa and Hamilton communities.

**Table 4:** Correlations between midpoint avg. of N1-N2 copies/copies PMMoV and laboratory positive cases, hospital admissions, ICU admissions, and deaths in Ottawa and Hamilton, respectively, post significant natural and vaccination immunization and during the onset of the Omicron BA.1 (B.1.1.529.1) VOC.

| Ottawa |  | 5-day midpoint avg. laboratory positive cases | 5-day midpoint avg. hospital admissions | 5-day midpoint avg. ICU admissions | 5-day midpoint avg. deaths |
| --- | --- | --- | --- | --- | --- |
| | Lead time ($\Delta t$) | 0d | 4d | 14d | 18d |
|  | Pearson's R | 0.861 | 0.947 | 0.911 | 0.905 |
| Hamilton |  | 3-day midpoint avg. laboratory positive cases | 3-day midpoint avg. hospital admissions | 3-day midpoint avg. ICU admissions | 3-day midpoint avg. deaths |
| | Lead time ($\Delta t$) | 0d | 3d | 2d | 9d |
|  | Pearson's R | 0.727 | 0.837 | 0.673 | 0.663 |

**Figure 3:**
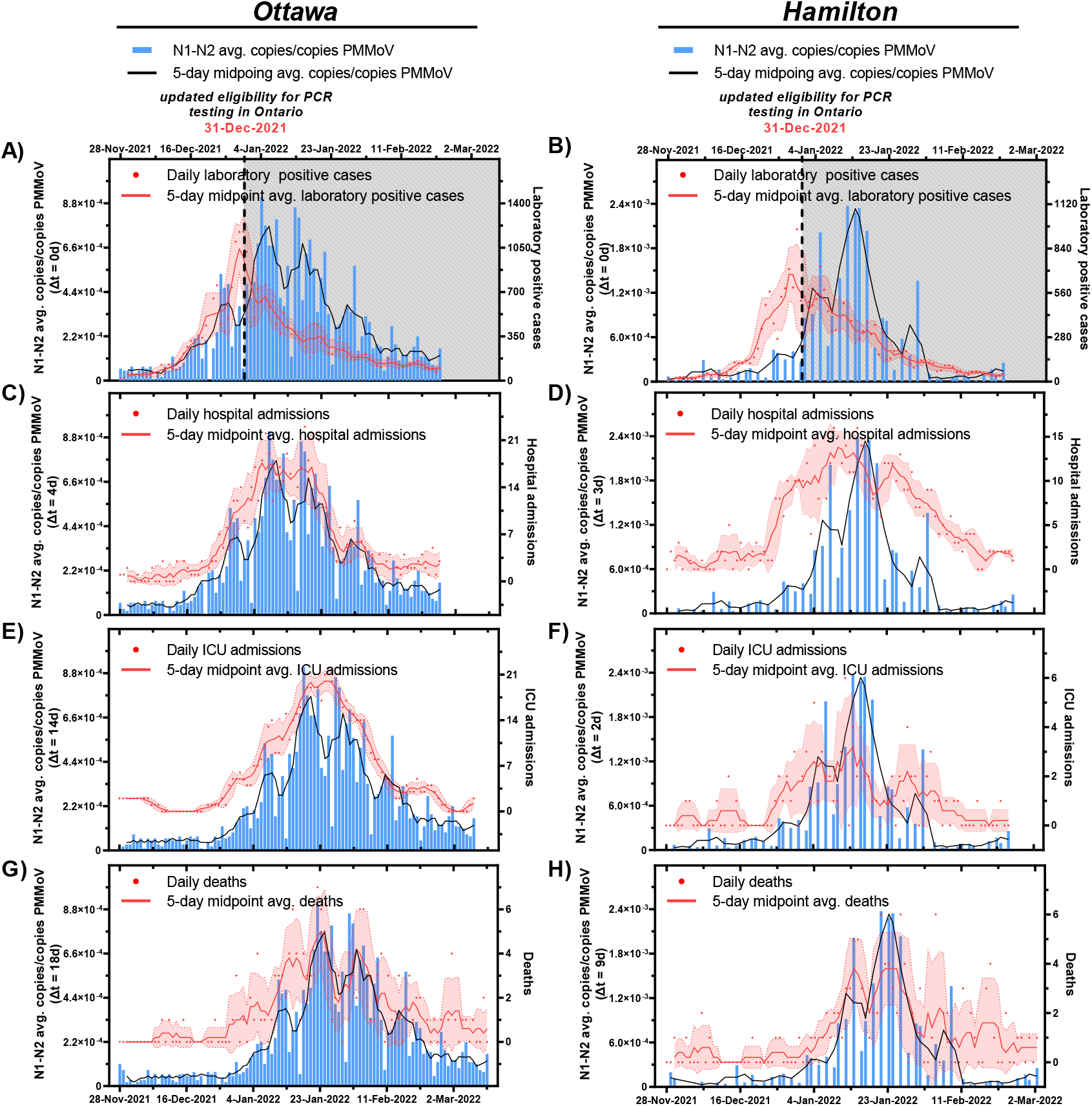
Relation between SARS-CoV-2 wastewater signal (advanced by respective “Δt” time lag on the x-axis) and A) clinical COVID-19 positive cases in Ottawa and B) clinical COVID-19 positive cases in Hamilton, C) hospital admissions in Ottawa and D) hospitalization admissions in Hamilton, E) ICU admissions in Ottawa and F) ICU admissions in Hamilton, and G) COVID-19 caused deaths in Ottawa, and H) COVID-19 caused deaths in Hamilton during the Omicron BA.1 (B.1.1.529.1) surge (Nov. 28^th^, 2021, to Feb. 22^nd^, 2022). *\*Laboratory positive cases in Ottawa and Hamilton are underreported due to updated PCR eligibility in Ontario as of December 31*^*st*^, *2021*

### 3.4 Relation between WWS signal and epidemiological metrics following gained natural immunity via recent surge, Omicron BA.2 (B.1.1.529.2) post protection surge

A facemask mandate and other public health protections such as gathering limits, and proof vaccination requirements were present throughout the entirety of the studied period until March 21^st^, 2022, when facemasks were no longer required in most indoor public settings (except for public transit, health care settings, long-term care homes, and congregate settings) (Government of Ontario, 2022). Facemasks were proven to provide the highest level of personal protection against contracting COVID-19 in indoor public settings (Andrejko et al., 2022). However, immediately following the removal of facemask mandates on Mar. 21^st^, 2022, a steep rise in the PMMoV-normalized SARS-CoV-2 viral signal measurements was observed in both Ottawa and Hamilton as the Omicron BA.1 (B.1.1.529.1) VOC resurges, along with a concurring surge of the highly transmissible Omicron BA.2 (B.1.1.529.2) subvariant. As of March 27^th^, 2022, 50% of laboratory positive cases were reportedly infected with the Omicron BA.2 (B.1.1.529.2) subvariant (Figure S2 in Supplemental Material) in Ottawa. During the Omicron BA.2 (B.1.1.529.2) surge, the 5-day average PMMoV-normalized SARS-CoV-2 measurement reached the highest recorded peak throughout the entirety of the study period after the previous highest peak measured during the Alpha (B.1.1.7) surge in both Ottawa and Hamilton (Figure 1). Unusually during this surge, however, all the four studied epidemiological metrics (laboratory positive cases, hospital admissions, ICU admissions, and deaths) exhibited lower values compared to those recorded during the preceding Alpha (B.1.1.7) and Omicron BA.1 (B.1.1.529.1) surges in both communities (Figure 1), even though evidence suggests a similar risk for individual infected with the Omicron BA.1 and BA.2 subvariants to be hospitalized and develop severe COVID-19 symptoms (Wolter et al., 2022). This observation is dissimilar to those made during the previous surges where all the clinical metrics reached peaks that were proportional to the wastewater measurements during the preceding wildtype, Alpha (B.1.1.7), Delta (B.1.617.2), and Omicron BA.1 (B.1.1.529.1) surges (Figure 1). This is likely attributed to enhanced natural immunization in the most frail members of the community (the elderly and immunocompromised) due to reinfection (at least 90 days after first infection) from the Omicron BA.1 (B.1.1.529.1) surge (Abu-Raddad et al., 2021), and enhanced vaccination immunization from receiving a booster COVID-19 vaccine dose which provides up to 90% protection against severe or fatal disease by the Omicron BA.2 (B.1.1.529.2) VOC at 7 weeks since reception of a third COVID-19 mRNA vaccine dose (Iorio A. et al., 2022); resulting in a lower hospitalization and fatality outcomes throughout the course of the Omicron BA.2 (B.1.1.529.2) surge.

A moderate positive correlation between PMMoV-normalized SARS-CoV-2 viral signal and laboratory positive cases was observed in Ottawa at Δt = 0d (R = 0.579), while a stronger correlation was observed at Δt = 0d (R = 0.847) in Hamilton (Table 5). It is noteworthy that during this period, PCR eligibility in Ontario was still limited to only symptomatic and high-risk individuals. It is thus very likely that the reported laboratory positive cases in both studied communities are underreported, and a stronger correlation could have been observed as shown previously during the Omicron BA.1 (B.1.1.529.1) surge before Dec. 31^st^, 2021 (Table 4, Figure 3 A and B). According to personal communications, personnel from a high-risk occupational setting who were still eligible for PCR testing of COVID-19 in Ontario post-Dec. 31^st^, 2021, reported a higher magnitude of positive COVID-19 test results during the Omicron BA.2 (B.1.1.529.2) compared to the previous Omicron BA.1 (B.1.1.529.1) surge. The trends displayed by those institutions displayed a similar visual trend with the wastewater viral signal with a lead time of approximately 5 days from the PMMoV-normalized viral signal measured in wastewater, further suggesting an underestimation of the true population-wide laboratory positive cases in both Ottawa and Hamilton. This further aligns with evidence of higher transmissibility of the Omicron BA.2 (B.1.1.529.2) sub-lineage compared to the preceding BA.1 (B.1.1.529.1) sub-lineage (Lyngse et al., 2022). Therefore, even as clinical testing eligibility was restricted and testing results are less representative of community incidence, wastewater viral signal measurement likely remained an indicator of incidence during the Omicron BA.2 (B.1.1.529.2) surge.

**Table 5:** Correlations between midpoint avg. of N1-N2 copies/copies PMMoV and laboratory positive cases, hospital admissions, ICU admissions, and deaths in Ottawa and Hamilton, respectively, post significant natural and vaccination immunization and during the onset of the Omicron BA.2 (B.1.1.529.2) VOC.

| Over 70% of the total population fully vaccinated Mar. 21<sup>st</sup>, 2022 – May 26<sup>th</sup>, 2022, 2022, Omicron surge |  |  |  |  |  |
| --- | --- | --- | --- | --- | --- |
| Ottawa |  | 5-day midpoint avg. laboratory positive cases | 5-day midpoint avg. hospital admissions | 5-day midpoint avg. ICU admissions | 5-day midpoint avg. deaths |
| | Lead time ($\Delta t$) | 0d | 5d | 14d | 18d |
|  | Pearson's R | 0.579 | 0.784 | 0.557 | 0.356 |

| Hamilton |  | 3-day midpoint avg.<br>laboratory positive<br>cases | 3-day midpoint avg.<br>hospital admissions | 3-day midpoint avg.<br>ICU admissions | 3-day midpoint avg.<br>deaths |
| --- | --- | --- | --- | --- | --- |
| | Lead time ($\Delta t$) | 0d | 3d | 2d | 16d |
|  | Pearson's R | 0.847 | 0.612 | 0.445 | 0.252 |

Findings from the previous wildtype, Alpha (B.1.1.7), Delta (B.1.617.2), and the first Omicron BA.1 (B.1.1.529.1) surges have suggested that wastewater was a reliable indicator of disease burden based on its relationship with the clinical metrics of hospitalization, ICU admissions, and deaths. During the second Omicron BA.2 (B.1.1.529.2) surge, although the PMMoV-normalized SARS-CoV-2 viral signal measurements displayed strong correlations with new hospital admissions at Δt = 5d in Ottawa (R = 0.784) and at Δt = 3d in Hamilton (R = 0.612) (Table 5), they are notably weaker compared to the correlations from the previous Omicron BA.1 (B.1.1.529.1) surge (R = 0.947 at Δt = 4d in Ottawa and R = 0.837 at Δt = 3d in Hamilton) prior to the removal of facemask mandate in both communities (Table 4). This is likely because hospitalizations during this Omicron BA.2 (B.1.1.529.2) surge fluctuate more irregularly at lower counts (between 0 and 11) (Figure 5 C and D) compared to the previous Omicron BA.1 (B.1.1.529.1) surge where hospital admissions ranged from 0 to 22 (Figure 4 C and D). Furthermore, a moderate positive correlation exists between the PMMoV-normalized viral signal measurements in wastewater and ICU admissions at Δt = 14d in Ottawa (R = 0.557) and at Δt = 2d in Hamilton (R = 0.445) (Table 5). Lastly, a weak positive correlation was observed between the PMMoV-normalized viral signal and COVID-19-related deaths at Δt = 18d in Ottawa (R = 0.356) and at Δt = 16d in Hamilton (R = 0.252) (Table 5). COVID-19 caused deaths during the Omicron BA.2 (B.1.1.529.2) surge does not visually exhibit a notable curve-like trend with increasing wastewater viral signal in both Ottawa (Figure 4 G) and Hamilton (Figure 4 H), unlike what was been previously observed with preceding VOC surges prior to Mar. 21^st^, 2022 (Figure 1). Compared to the previous Omicron BA.1 (B.1.1.529.1) surge, not only are the correlations weaker between the clinical metrics indicative of disease burden (hospitalization, ICU admissions, and deaths) and wastewater viral signal during the Omicron BA.2 (B.1.1.529.2) surge, but the magnitudes of all the three metrics are reportedly lower (Figure 1). It is hypothesized that the lower hospitalizations, ICU admissions, and deaths during the Omicron BA.2 (B.1.1.529.2) surge are attributed to enhanced natural immunization and enhanced vaccination immunization against severe disease outcomes. Another contributor to higher hospitalizations between Dec. 2021 and Feb. 2022 during the Omicron BA.1 (B.1.1.529.1) surge compared to Mar. 2022 onwards during the Omicron BA.2 (B.1.1.529.2) surge could be a form of morbidity or mortality displacement in which the most frail members of the population are affected by the first Omicron BA.1 (B.1.1.529.1) surge. Due to reduced protection posed by Omicron’s vaccine-escape capabilities, the most frail people in the community (the elderly, immunocompromised) who were protected from previous VOCs may have had severe COVID-19-related complications (hospitalization, ICU admission, or death) early in the Omicron BA.1 (B.1.1.529.1) surge. During the onset of the Omicron BA.2 (B.1.1.529.2) from Mar. 2022 onwards, those frail members of the community have either succumbed to the illness or had recovered leaving a less-frail population for which significant natural and vaccination immunization may still convey protection from severe outcomes. Therefore, during the surge of a less virulent VOC in a community with significant natural and vaccination immunization, wastewater is shown to be less of an indicator of disease burden.

**Figure 4:**
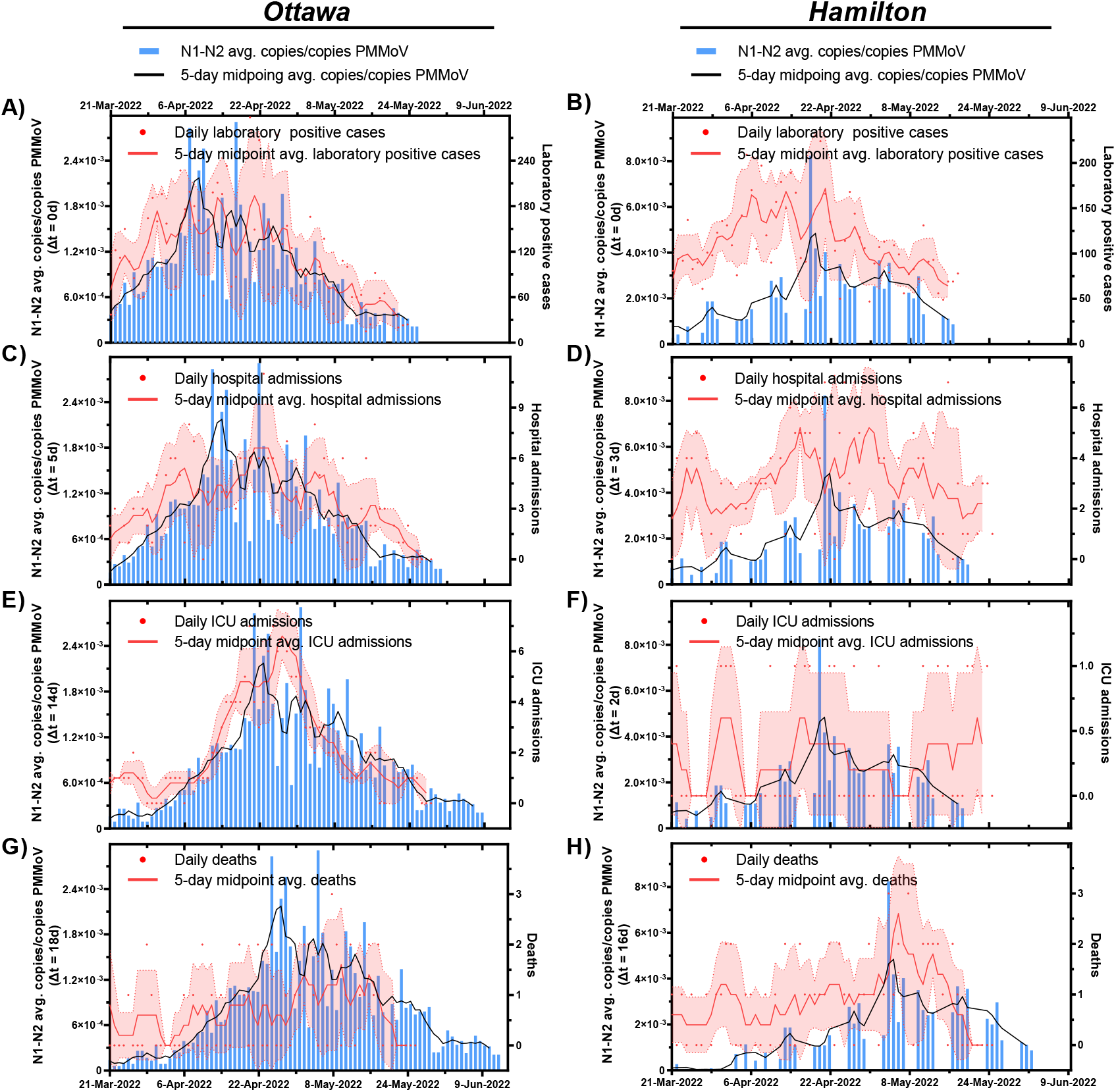
Relation between SARS-CoV-2 wastewater signal (advanced by respective “Δt” time lag on the x-axis and A) clinical COVID-19 positive cases in Ottawa and B) clinical COVID-19 positive cases in Hamilton, C) hospital admissions in Ottawa and D) hospitalization admissions in Hamilton, E) ICU admissions in Ottawa and F) ICU admissions in Hamilton, and G) COVID-19 caused deaths in Ottawa, and H) COVID-19 caused deaths in Hamilton during the Omicron BA.2 (B.1.1.529.2) surge post removal of protection/face masking protection (Mar. 21^th^, 2022 to May 26^th^, 2022). *\*Laboratory positive cases in Ottawa and Hamilton are underreported due to updated PCR eligibility in Ontario as of December 31*^*st*^, *2021*

**Figure 5:**
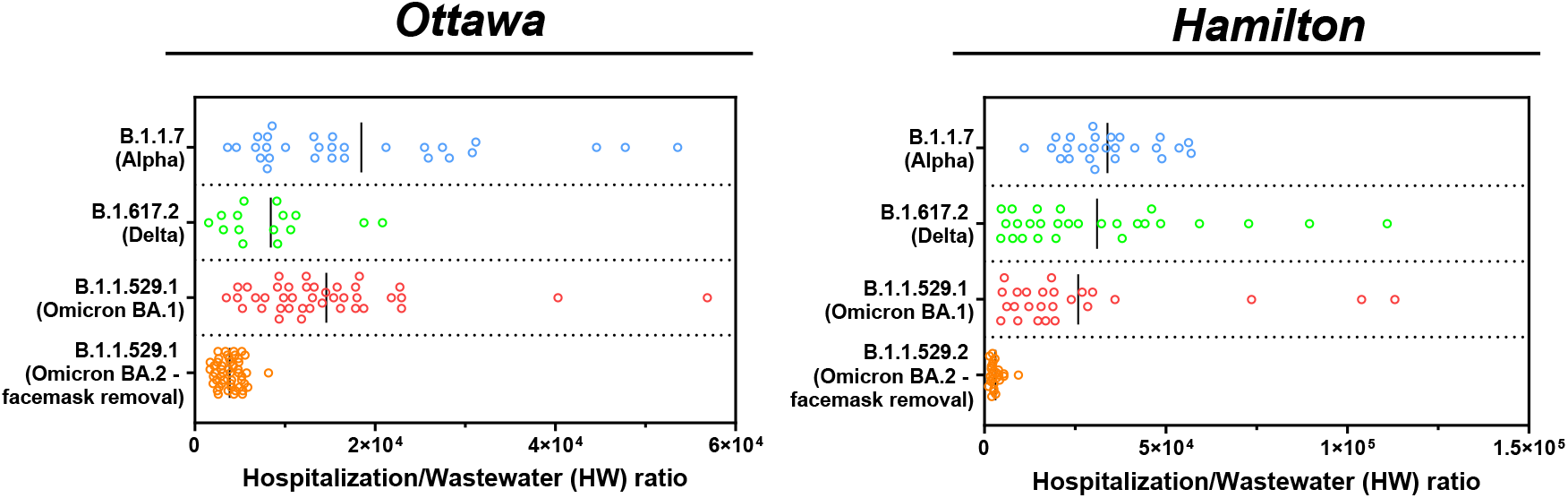
Comparison of hospitalization to wastewater ratio amplitude during the onset of the B.1.1.7 (Alpha), B.1.617 (Delta), B.1.1.529.1 (Omicron BA.1), and B.1.1.529.2 (Omicron BA.2 – post facemask removal) VOCs in Ottawa and Hamilton.

Post significant vaccination immunization, between the past 3 – 4 months of authoring this article (Jun. 2022), the link to disease incidence was weakened, but the link to hospitalizations was not. During the post protection Omicron BA.2 (B.1.1.529.2) surge, wastewater is shown to be an indicator of incidence and infections of a less virulent VOC, and less of an indicator of disease burden either due to significant natural and vaccination immunization or due to morbidity displacement.

### 3.5 Progression of hospitalization data as an indicator of disease virulence

Wastewater signal as a predictor and indicator of community disease burden is demonstrated during wildtype COVID-19 and the spread of the B.1.1.7 (Alpha), B.1.617.2 (Delta), and B.1.1.529.1 (Omicron BA.1 – prior to protection removal) VOCs, but not during the B.1.1.529.2 (Omicron BA.2 – post protection/facemask removal). During the onset of the wildtype COVID-19 between Mar. 2020 and Feb. 2021, wastewater viral signal was a predictor and indicator of both incidence (laboratory positive cases) and disease burden (hospital admissions) in both Ottawa (Figure 1 A and C) and Hamilton (Figure 1 B and D). The B.1.1.7 (Alpha) VOC further caused infectious surges of COVID-19 disease between Mar. and May 2021 while vaccine rollout was still limited and only 2.2% and 2.7% of the Ottawa and Hamilton population were fully vaccinated against COVID-19 (2 approved vaccine doses) by Mar. 23^rd^, 2021 (onset of B.1.1.7 (Alpha) VOC), respectively (Public Health Ontario, 2022a). With the majority of the population in both communities lacking natural and vaccination immunity, the B.1.1.7 (Alpha) surge had the highest morbidity outcome (i.e., the highest counts of hospital admissions, ICU admissions, and deaths due to COVID-19 complications) throughout the studied period (Figure 1). The elevated magnitude of the corresponding hospitalization-to-wastewater (HW) ratio is further indicative of the virulence of the B.1.1.7 (Alpha) VOC in both communities (Figure 5). In the course of the wildtype COVID-19 and the B.1.1.7 (Alpha) VOC surge, the wastewater signal strongly correlated to both COVID-19 positive clinical cases and hospital admissions in Ottawa and Hamilton (Figure 1). Thus, prior to significant vaccination immunization, wastewater functioned as an effective predictor and indicator of both incidence (laboratory positive cases) and disease burden (hospital admissions) with virulent dominant VOC.

Although vaccine rollout was limited during the onset of the B.1.1.7 (Alpha) VOC, vaccination rates increased rapidly from Jun. 2021 through Aug. 2021 concurrently as the more transmissible B.1.617.2 (Delta) VOC surged through the cities. By 23^rd^ Jul. 23^rd^/Aug. 5^th^, 2021, prior to the onset of the B.1.617.2 (Delta) VOC, at least 60% of the total population in Ottawa/Hamilton were fully immunized (2 approved vaccine doses) and by Aug. 29^th^/Oct. 9^th^, 2021, in Ottawa/Hamilton, at least 70% of the total population were fully immunized (Public Health Ontario, 2022a). With significant vaccination immunization achieved in both cities not long from the onset of the B.1.617.2 (Delta) VOC, both communities benefited from peak vaccination immunity. There was further the benefit of enhanced natural immunity for recovered individuals previously infected with the B.1.1.7 (Alpha) VOC during its dominance. Consequently, both studied communities experienced the lowest disease incidence (Figure 1 A and B) and hospital admissions (Figure 1 C and D) throughout the entire studied period during the B.1.617.2 (Delta) VOC surge. With lower proportions of symptomatic infections occurring in the general population during the B.1.617.2 (Delta) VOC surge, the link between wastewater signal and COVID-19 incidence was weakened (population-wide PCR testing in Ontario was available during this period) (Figure 2 A and B) while the relation to disease hospitalization was preserved (Figure 2 C and D). While the lowest hospitalization counts and wastewater signals were experienced in both communities during the dominance of the B.1.617.2 (Delta) VOC, an elevated HW ratio is still observed, reflecting the virulent nature of the B.1.617.2 (Delta) VOC (Figure 5).

In December 2021, the highly infectious B.1.1.529.1 (Omicron BA.1) VOC became dominant in both studied communities. By then, there is a possibility of waning vaccination protection as over 20 weeks have passed since ≥ 70% of the total population in both communities achieved full vaccination immunization, along with the vaccine-escape qualities of the B.1.1.529.1 (Omicron BA.1) VOC. These factors have likely contributed to the highest surge in laboratory positive cases (Figure 1 A and B) with the second-highest morbid outcomes (i.e. second highest peaks in hospital admissions, ICU admissions, and COVID-19 related deaths after the B.1.1.7 (Alpha) VOC) in Ottawa (Figure 1 C, E and G) and Hamilton (Figure 1 D, F, and H). The virulence of this VOC is further reflected by an elevated HW ratio (Figure 5), similarly to the preceding VOCs. During this period (Dec. 2021 – Feb. 2022), wastewater viral signal measurements likely have a strong correlation to incidence (by Dec. 31^st^, 2021, laboratory positive cases in both communities are underreported due to province-wide updated PCR eligibility) and once again maintained a strong correlation to hospitalization, albeit with shorter lead times of 4 days (Ottawa) and 3 days (Hamilton) (Figure 3 C and D), which are likely attributed to the shorter incubation time of the B.1.1.529.1 (Omicron BA.1) VOC compared to the preceding VOCs.

In mid-March 2022, the highly infections B.1.1.529.2 (Omicron BA.2) VOC predominant the latest surge in Ottawa and Hamilton (Figure 1 & Figure 4). By early Feb. 2022/early Mar. 2022 (prior to the onset of the B.1.1.529.2 (Omicron BA.2)), 50% of the total population received a booster dose (third dose of approved COVID-19 vaccine) in Ottawa/Hamilton (Public Health Ontario, 2022a); increasing vaccine-induced effectiveness for both communities against the B.1.1.529.1 & B.1.1.529.2 (Omicron BA.1 & BA.2) VOC. Natural immunity is further heightened due to SARS-CoV-2 infections during the previous B.1.1.529.1 (Omicron BA.1). As such, the populations are at peak vaccination and natural immunity during the B.1.1.529.2 (Omicron BA.2); a similar scenario to the B.1.617.2 (Delta) VOC surge. Concurrently, the mandatory facemask requirement in most public indoor spaces was removed province wide as of Mar. 21^st^, 2022, whilst the B.1.1.529.2 (Omicron BA.2) VOC began to spread through the cities between Mar. 2022 through May 2022. Lower morbidity outcomes were observed during the B.1.1.529.2 (Omicron BA.2) VOC despite being of similar severity to the B.1.1.529.1 (Omicron BA.1) VOC. This is further reflected by a lower magnitude of HW ratio in both Ottawa and Hamilton during the B.1.1.529 (Omicron BA.2) surge, indicative of less virulent dominant VOC during a period of peak vaccination and natural immunization (Figure 5). During this surge, wastewater relation to the clinical metrics is expected to be similar to that observed during the B.1.617.2 (Delta) surge due to being of similar immunization dynamics. However, the removal of facemask protection during the second Omicron surge has resulted in a rapid surge in laboratory positive cases (albeit being underreported due to limited population-wide PCR testing), hence strongly correlating to the wastewater signal in Ottawa (Figure 4 A) and Hamilton (Figure 4 B). The wastewater signal is also displayed as a strong indicator of hospital admissions in both communities (Figure 4 C and D). The evolution of the relation between wastewater and community disease incidence and burden throughout the studied period of the COVID-19 pandemic is summarized in Table 6.

**Table 6:**
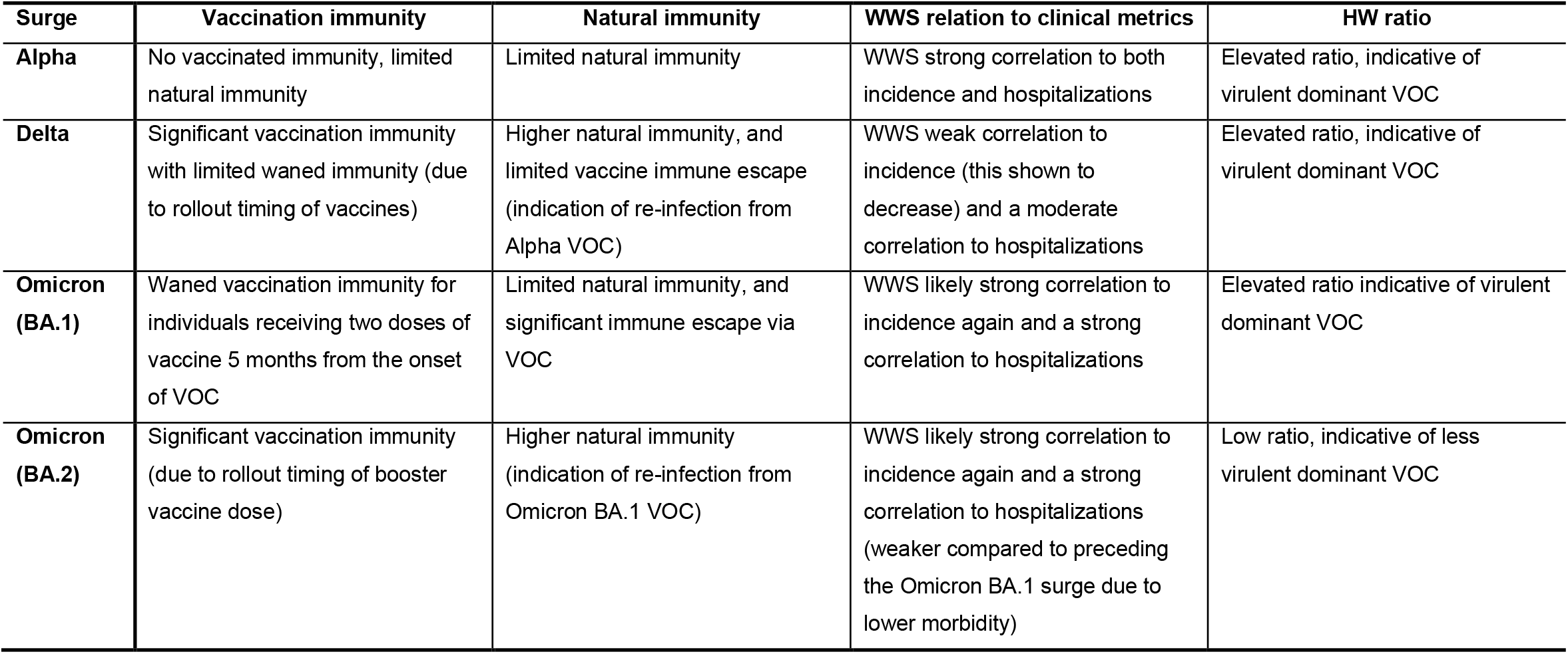
Evolution of WWS relation to clinical epidemiological metrics throughout the studied period of the COVID-19 pandemic

## 4 Conclusions

This study demonstrates the progression of WWS of SASR-CoV-2 in wastewater as an indicator of disease incidence and disease burden in various stages of the pandemic. During periods with no vaccinated or natural immunity when the wildtype COVID-19 and the Alpha (B.1.1.7) VOC predominated, wastewater is a strong predictor and indicator for both disease incidence and burden. The high virulence morbidity outcomes from the Alpha (B.1.1.7) VOC were reflected by an elevated HW ratio. Further into the pandemic, during periods with peak vaccination immunization (2 – 4 weeks prior after receiving two approved vaccine doses prior to vaccine waning) when the Delta (B.1.617.2) VOC predominated, wastewater weakened as an indictor of laboratory positive cases, but it is a strong indicator for disease burden. Despite the Delta (B.1.617.2) VOC being characterized as more virulent and transmissible compared to the Alpha (B.1.1.7) VOC, the studied communities experienced the lowest morbidity outcome; yet the elevated HW ratio during the onset of this surge is indicative of the virulence of the Delta (B.1.6172) VOC. During the periods of waned vaccination immunization (similar scenario to the period during the Alpha surge) during the first Omicron BA.1 (B.1.1.529.1) surge, wastewater is a strong indicator for both disease incidence and burden. The high morbidity outcome experienced by the studied communities was reflected by the HW ratio during this surge. Lastly, returning to a period of peak natural immunity (due to reinfection) and peak vaccination immunization (due to booster vaccine dose rollout) achieved 3 weeks prior to the Omicron BA.2 (B.1.1.529.2) surge wastewater is a strong indicator for both disease incidence and hospital admissions. While both the Omicron BA.1 and BA.2 are characterized with similar virulence, the studied communities experienced lower morbidity outcomes during the Omicron BA.2 surge due to significant natural and vaccination immunization. This is further reflected by a low HW ratio. In the future, in the case that individuals would require seasonal vaccination for COVID-19 such that vaccination waning becomes limited, and while the vaccine is effective for the dominant VOC, WWS is expected to show moderate indication of incidence and strong indication of disease burden in the community. The wastewater to hospitalization data is also a good indicator of virulence when widespread clinical testing is limited.

## Data Availability

All data produced in the present study are available upon reasonable request to the authors

## Acknowledgements

The authors wish to acknowledge the help and assistance of the University of Ottawa, the Ottawa Hospital, the Children’s Hospital of Eastern Ontario, the Children’s Hospital of Eastern Ontario’s Research Institute, Ottawa Public Health, City of Hamilton, Public Health Hamilton, Public Health Ontario, and all of their employees who were involved in this project. In particular, we would like to acknowledge the contributions of Cameron McDermaid and Monir Taha of Ottawa Public Health. Their time, facilities, resources, and feedback are greatly appreciated.

## References

Abu-Raddad, L.J., Chemaitelly, H., Bertollini, R., 2021. Severity of SARS-CoV-2 Reinfections as Compared with Primary Infections. N. Engl. J. Med. 385, 2487–2489. https://doi.org/10.1056/NEJMc2108120

Ahmed, W., Angel, N., Edson, J., Bibby, K., Bivins, A., O’Brien, J.W., Choi, P.M., Kitajima, M., Simpson, S.L., Li, J., Tscharke, B., Verhagen, R., Smith, W.J.M., Zaugg, J., Dierens, L., Hugenholtz, P., Thomas, K. V., Mueller, J.F., 2020. First confirmed detection of SARS-CoV-2 in untreated wastewater in Australia: A proof of concept for the wastewater surveillance of COVID-19 in the community. Sci. Total Environ. 728, 138764. https://doi.org/10.1016/j.scitotenv.2020.138764

Andrejko, K.L., Pry, J.M., Myers, J.F., Fukui, N., DeGuzman, J.L., Openshaw, J., Watt, J.P., Lewnard, J.A., Jain, S., Team, C.C.-19 C.-C.S., Team, C.C.-19 C.-C.S., Abdulrahim, Y., Barbaduomo, C.M., Bermejo, M.I., Cheunkarndee, J., Cornejo, A.F., Corredor, S., Dabbagh, N., Dong, Z.N., Dyke, A., Fang, A.T., Felipe, D., Frost, P.M., Ho, T., Javadi, M.H., Kaur, A., Lam, A., Li, S.S., Miller, M., Ni, J., Park, H., Poindexter, D.J., Samani, H., Saretha, S., Spencer, M., Spinosa, M.M., Tran, V.H., Walas, N., Wan, C., Xavier, E., 2022. Effectiveness of Face Mask or Respirator Use in Indoor Public Settings for Prevention of SARS-CoV-2 Infection — California, February–December 2021. Morb. Mortal. Wkly. Rep. 71, 212. https://doi.org/10.15585/MMWR.MM7106E1

Andrews, N., Stowe, J., Kirsebom, F., Toffa, S., Rickeard, T., Gallagher, E., Gower, C., Kall, M., Groves, N., O’Connell, A.-M., Simons, D., Blomquist, P.B., Zaidi, A., Nash, S., Aziz, N.I.B.A., Thelwall, S., Dabrera, G., Myers, R., Amirthalingam, G., Gharbia, S., Barrett, J.C., Elson, R., Ladhani, S.N., Ferguson, N., Zambon, M., Campbell, C.N.J., Brown, K., Hopkins, S., Chand, M., Ramsay, M., Bernal, J.L., 2022. Covid-19 Vaccine Effectiveness against the Omicron (B.1.1.529) Variant. N. Engl. J. Med. 386, 1532–1546. https://doi.org/10.1056/NEJMOA2119451

Asghar, H., Diop, O.M., Weldegebriel, G., Malik, F., Shetty, S., El Bassioni, L., Akande, A.O., Al Maamoun, E., Zaidi, S., Adeniji, A.J., Burns, C.C., Deshpande, J., Oberste, M.S., Lowther, S.A., 2014. Environmental Surveillance for Polioviruses in the Global Polio Eradication Initiative. J. Infect. Dis. 210, S294–S303. https://doi.org/10.1093/INFDIS/JIU384

Backer, J.A., Eggink, D., Andeweg, S.P., Veldhuijzen, I.K., Maarseveen, N. van, Vermaas, K., Vlaemynck, B., Schepers, R., Hof S. van den, Reusken, C.B.E.., Wallinga, J., 2022. Shorter serial intervals in SARS-CoV-2 cases with Omicron BA.1 variant compared to Delta variant in the Netherlands, 13 -26 December 2021. med 2003–2005. https://doi.org/10.1101/2022.01.18.22269217

Bar-Or, I., Yaniv, K., Shagan, M., Ozer, E., Erster, O., Mendelson, E., Mannasse, B., Shirazi, R., Kramarsky-Winter, E., Nir, O., Abu-Ali, H., Ronen, Z., Rinott, E., Lewis, Y.E., Friedler, E., Bitkover, E., Paitan, Y., Berchenko, Y., Kushmaro, A., 2020. Regressing SARS-CoV-2 sewage measurements onto COVID-19 burden in the population: A proof-of-concept for quantitative environmental surveillance. medRxiv 1–11. https://doi.org/10.1101/2020.04.26.20073569

City of Hamilton, 2022. Status of Cases in Hamilton | City of Hamilton, Ontario, Canada [WWW Document]. URL https://www.hamilton.ca/coronavirus/status-cases-in-hamilton (accessed 1.17.22).

COVIDPoops19, 2022. Summary of global SARS-CoV-2 wastewater monitoring efforts by UC Merced researchers [WWW Document]. Univ. Calif. Merced. URL https://ucmerced.maps.arcgis.com/apps/dashboards/c778145ea5bb4daeb58d31afee389082 (accessed 6.6.22).

D’Aoust, P.M., Graber, T.E., Mercier, E., Montpetit, D., Alexandrov, I., Neault, N., Baig, A.T., Mayne, J., Zhang, X., Alain, T., Servos, M.R., Srikanthan, N., MacKenzie, M., Figeys, D., Manuel, D., Jüni, P., MacKenzie, A.E., Delatolla, R., 2021a. Catching a resurgence: Increase in SARS-CoV-2 viral RNA identified in wastewater 48 h before COVID-19 clinical tests and 96 h before hospitalizations. Sci. Total Environ. 770. https://doi.org/10.1016/j.scitotenv.2021.145319

D’Aoust, P.M., Mercier, E., Montpetit, D., Jia, J.J., Alexandrov, I., Neault, N., Baig, A.T., Mayne, J., Zhang, X., Alain, T., Langlois, M.A., Servos, M.R., MacKenzie, M., Figeys, D., MacKenzie, A.E., Graber, T.E., Delatolla, R., 2021b. Quantitative analysis of SARS-CoV-2 RNA from wastewater solids in communities with low COVID-19 incidence and prevalence. Water Res. 188, 116560. https://doi.org/10.1016/j.watres.2020.116560

D’Aoust, P.M., Tian, X., Tasneem, S., Xiao, A., Mercier, E., 2022. Wastewater to clinical case (WC) ratio of COVID-19 identifies insufficient clinical testing, onset of new variants of concern and population immunity in urban communities. medRxiv. https://doi.org/10.1101/2022.04.19.22274052

D’Aoust, P.M., Towhid, S.T., Mercier, É., Hegazy, N., Tian, X., Bhatnagar, K., Zhang, Z., Mackenzie, A.E., Graber, T.E., Delatolla, R., 2021c. COVID-19 monitoring in rural communities: First comparison of lagoon and pumping station samples for wastewater-based epidemiology. Environ. Sci. Water Res. Technol. https://doi.org/10.1016/j.scitotenv.2021.149618

Galani, A., Aalizadeh, R., Kostakis, M., Markou, A., Alygizakis, N., Lytras, T., Adamopoulos, P.G., Peccia, J., Thompson, D.C., Kontou, A., Karagiannidis, A., Lianidou, E.S., Avgeris, M., Paraskevis, D., Tsiodras, S., Scorilas, A., Vasiliou, V., Dimopoulos, M.A., Thomaidis, N.S., 2022. SARS-CoV-2 wastewater surveillance data can predict hospitalizations and ICU admissions. Sci. Total Environ. 804, 150151. https://doi.org/10.1016/J.SCITOTENV.2021.150151

Gerrity, D., Papp, K., Stoker, M., Sims, A., Frehner, W., 2021. Early-pandemic wastewater surveillance of SARS-CoV-2 in Southern Nevada: Methodology, occurrence, and incidence/prevalence considerations. Water Res. X 10, 100086. https://doi.org/10.1016/j.wroa.2020.100086

Gonzalez, R., Curtis, K., Bivins, A., Bibby, K., Weir, M.H., Yetka, K., Thompson, H., Keeling, D., Mitchell, J., Gonzalez, D., 2020. COVID-19 surveillance in Southeastern Virginia using wastewater-based epidemiology. Water Res. 186, 116296. https://doi.org/10.1016/j.watres.2020.116296

Government of Canada, 2022. COVID-19 vaccination coverage in Canada [WWW Document]. URL https://health-infobase.canada.ca/covid-19/vaccination-coverage/ (accessed 6.1.22).

Government of Ontario, 2022. Statement from Ontario’s Chief Medical Officer of Health | Ontario Newsroom [WWW Document]. URL https://news.ontario.ca/en/statement/1001732/statement-from-ontarios-chief-medical-officer-of-health (accessed 5.18.22).

Government of Ontario, 2021. Updated Eligibility for PCR Testing and Case and Contact Management Guidance in Ontario | Ontario Newsroom [WWW Document]. URL https://news.ontario.ca/en/backgrounder/1001387/updated-eligibility-for-pcr-testing-and-case-and-contact-management-guidance-in-ontario (accessed 1.16.22).

Graham, K.E., Loeb, S.K., Wolfe, M.K., Catoe, D., Sinnott-Armstrong, N., Kim, S., Yamahara, K.M., Sassoubre, L.M., Mendoza Grijalva, L.M., Roldan-Hernandez, L., Langenfeld, K., Wigginton, K.R., Boehm, A.B., 2021. SARS-CoV-2 RNA in Wastewater Settled Solids Is Associated with COVID-19 Cases in a Large Urban Sewershed. Environ. Sci. Technol. 55, 488–498. https://doi.org/10.1021/acs.est.0c06191

Haramoto, E., Malla, B., Thakali, O., Kitajima, M., 2020. First environmental surveillance for the presence of SARS-CoV-2 RNA in wastewater and river water in Japan. Sci. Total Environ. 737, 140405. https://doi.org/10.1016/J.SCITOTENV.2020.140405

Hellmér, M., Paxéus, N., Magnius, L., Enache, L., Arnholm, B., Johansson, A., Bergström, T., Norder, H., 2014. Detection of pathogenic viruses in sewage provided early warnings of hepatitis A virus and norovirus outbreaks. Appl. Environ. Microbiol. 80, 6771–6781. https://doi.org/10.1128/AEM.01981-14

Hogan, A.B., Wu, S.L., Doohan, P., Watson, O.J., Winskill, P., Charles, G., Barnsley, G., Riley, E.M., Khoury, D.S., Ferguson, N.M., Ghani, A.C., 2021. Report 48: The value of vaccine booster doses to mitigate the global impact of the Omicron SARS-CoV-2 variant. https://doi.org/10.25561/93034

Inokuchi, R., Morita, K., Iwagami, M., Watanabe, T., Tamiya, N., 2021. Changes in the proportion and severity of patients with fever or common cold symptoms utilizing an after-hours house call medical service during the COVID-19 pandemic in Tokyo, Japan: a retrospective cohort study. BMC Emerg. Med. 21. https://doi.org/10.1186/s12873-021-00458-8

Iorio A., Little J., Linkins L., Abdelkader W., Bennett D., Lavis JN., 2022. COVID-19 Living Evidence Synthesis 6.31: What is the efficacy and effectiveness of available COVID-19 vaccines in general and specifically for variants of concern?

Kannan, S.R., Spratt, A.N., Cohen, A.R., Naqvi, S.H., Chand, H.S., Quinn, T.P., Lorson, C.L., Byrareddy, S.N., Singh, K., 2021. Evolutionary analysis of the Delta and Delta Plus variants of the SARS-CoV-2 viruses. J. Autoimmun. 124, 102715. https://doi.org/10.1016/J.JAUT.2021.102715

Kaplan, E.H., Wang, D., Wang, M., Malik, A.A., Zulli, A., Peccia, J., 2021. Aligning SARS-CoV-2 indicators via an epidemic model: application to hospital admissions and RNA detection in sewage sludge. Health Care Manag. Sci. 24, 320–329. https://doi.org/10.1007/s10729-020-09525-1

Kimball, A., Hatfield, K.M., Arons, M., James, A., Taylor, J., Spicer, K., Bardossy, A.C., Oakley, L.P., Tanwar, S., Chisty, Z., Bell, J.M., Methner, M., Harney, J., Jacobs, J.R., Carlson, C.M., McLaughlin, H.P, Stone, N., Clark, S., Brostrom-Smith, C., Page, L.C., Kay, M., Lewis, J., Russell, D., Hiatt, B., Gant, J., Duchin, J.S., Clark, T.A., Honein, M.A., Reddy, S.C., Jernigan, J.A., Baer, A., Barnard, L.M., Benoliel, E., Fagalde, M.S., Ferro, J., Smith, H.G., Gonzales, E., Hatley, N., Hatt, G., Hope, M., Huntington-Frazier, M., Kawakami, V., Lenahan, J.L., Lukoff, M.D., Maier, E.B., McKeirnan, S., Montgomery, P., Morgan, J.L., Mummert, L.A., Pogosjans, S., Riedo, F.X., Schwarcz, L., Smith, D., Stearns, S., Sykes, K.J., Whitney, H., Ali, H., Banks, M., Balajee, A., Chow, E.J., Cooper, B., Currie, D.W., Dyal, J., Healy, J., Hughes, M., McMichael, T.M., Nolen, L., Olson, C., Rao, A.K., Schmit, K., Schwartz, N.G., Tobolowsky, F., Zacks, R., Zane, S., 2020. Asymptomatic and Presymptomatic SARS-CoV-2 Infections in Residents of a Long-Term Care Skilled Nursing Facility — King County, Washington, March 2020. MMWR. Morb. Mortal. Wkly. Rep. 69, 377–381. https://doi.org/10.15585/MMWR.MM6913E1

Kocamemi, B.A., Kurt, H., Sait, A., Sarac, F., Saatci, A.M., Pakdemirli, B., 2020. SARS-CoV-2 Detection in Istanbul Wastewater Treatment Plant Sludges. medRxiv 2020.05.12.20099358. https://doi.org/10.1101/2020.05.12.20099358

La Rosa, G., Mancini, P., Bonanno Ferraro, G., Veneri, C., Iaconelli, M., Bonadonna, L., Lucentini, L., Suffredini, E., 2021. SARS-CoV-2 has been circulating in northern Italy since December 2019: Evidence from environmental monitoring. Sci. Total Environ. 750, 141711. https://doi.org/10.1016/j.scitotenv.2020.141711

Lee, J.J., Choe, Y.J., Jeong, H., Kim, M., Kim, S., Yoo, H., Park, K., Kim, C., Choi, S., Sim, J.W., Park, Y., Huh, I.S., Hong, G., Kim, M.Y., Song, J.S., Lee, J., Kim, E.J., Rhee, J.E., Kim, I.H., Gwack, J., Kim, Jungyeon, Jeon, J.H., Lee, W.G., Jeong, S., Kim, Jusim, Bae, B., Kim, J.E., Kim, H., Lee, H.Y., Lee, S.E., Kim, J.M., Park, H., Yu, M., Choi, J., Kim, Jia, Lee, H., Jang, E.J., Lim, D., Lee, S., Park, Y.J., 2021. Importation and Transmission of SARS-CoV-2 B.1.1.529 (Omicron) Variant of Concern in Korea, November 2021. J. Korean Med. Sci. 36, 2–5. https://doi.org/10.3346/JKMS.2021.36.E346

Levine-Tiefenbrun, M., Yelin, I., Katz, R., Herzel, E., Golan, Z., Schreiber, L., Wolf, T., Nadler, V., Ben-Tov, A., Kuint, J., Gazit, S., Patalon, T., Chodick, G., Kishony, R., 2021. Decreased SARS-CoV-2 viral load following vaccination. Nat. Med. 27, 790–792. https://doi.org/10.1038/s41591-021-01316-7

Lin, D.-Y., Gu, Y., Wheeler, B., Young, H., Holloway, S., Sunny, S.K., Moore, Z., Zeng, D., 2021. Effectiveness of Covid-19 Vaccines in the United States Over 9 Months: Surveillance Data from the State of North Carolina. medRxiv 2021.10.25.21265304. https://doi.org/10.1101/2021.10.25.21265304

Liu, L., Iketani, S., Guo, Y., Chan, J.F.W., Wang, M., Liu, Liyuan, Luo, Y., Chu, H., Huang, Yiming, Nair, M.S., Yu, J., Chik, K.K.H., Yuen, T.T.T., Yoon, C., To, K.K.W., Chen, H., Yin, M.T., Sobieszczyk, M.E., Huang, Yaoxing, Wang, H.H., Sheng, Z., Yuen, K.Y., Ho, D.D., 2022. Striking antibody evasion manifested by the Omicron variant of SARS-CoV-2. Nature 602, 676–681. https://doi.org/10.1038/S41586-021-04388-0

Liu, Y., Liu, J., Johnson, B.A., Xia, H., Ku, Z., Schindewolf, C., Widen, S.G., An, Z., Weaver, S.C., Menachery, V.D., Xie, X., Shi, P.-Y., 2022. Delta spike P681R mutation enhances SARS-CoV-2 fitness over Alpha variant. Cell Rep. 39, 110829. https://doi.org/10.1016/J.CELREP.2022.110829

Lyngse, F.P., Kirkeby, C.T., Denwood, M., Christiansen, L.E., Mølbak, K., Møller, C.H., Skov, R.L., Krause, T.G., Rasmussen, M., Sieber, R.N., Johannesen, T.B., Lillebaek, T., Fonager, J., Fomsgaard, A., Møller, F.T., Stegger, M., Overvad, M., Spiess, K., Mortensen, L.H., 2022. Transmission of SARS-CoV-2 Omicron VOC subvariants BA.1 and BA.2: Evidence from Danish Households. medRxiv 2022.01.28.22270044. https://doi.org/10.1101/2022.01.28.22270044

Mathieu, E., Ritchie, H., Ortiz-Ospina, E., Roser, M., Hasell, J., Appel, C., Giattino, C., Rodés-Guirao, L., 2021. A global database of COVID-19 vaccinations. Nat. Hum. Behav. 5, 947–953. https://doi.org/10.1038/S41562-021-01122-8

Medema, G., Heijnen, L., Elsinga, G., Italiaander, R., Brouwer, A., 2020. Presence of SARS-Coronavirus-2 RNA in Sewage and Correlation with Reported COVID-19 Prevalence in the Early Stage of the Epidemic in the Netherlands. Environ. Sci. Technol. Lett. 7, 511–516. https://doi.org/10.1021/acs.estlett.0c00357

Mizumoto, K., Kagaya, K., Zarebski, A., Chowell, G., 2020. Estimating the asymptomatic proportion of coronavirus disease 2019 (COVID-19) cases on board the Diamond Princess cruise ship, Yokohama, Japan, 2020. Eurosurveillance 25. https://doi.org/10.2807/1560-7917.ES.2020.25.10.2000180

Naughton, C.C., Roman, F.A., Grace Alvarado, A.F., Tariqi, A.Q., Deeming, M.A., Bibby, K., Bivins, A., Rose, J.B., Medema, G., Ahmed, W., Katsivelis, P., Allan, V., Sinclair, R., Zhang, Y., Kinyua, M.N., Author cnaughton, C., 2021. Show us the Data: Global COVID-19 Wastewater Monitoring Efforts, Equity, and Gaps. medRxiv 2021.03.14.21253564. https://doi.org/10.1101/2021.03.14.21253564

Nemudryi, A., Nemudraia, A., Wiegand, T., Surya, K., Buyukyoruk, M., Cicha, C., Vanderwood, K.K., Wilkinson, R., Wiedenheft, B., 2020. Temporal Detection and Phylogenetic Assessment of SARS-CoV-2 in Municipal Wastewater. Cell Reports Med. 1, 100098. https://doi.org/10.1016/J.XCRM.2020.100098

Ottawa Public Health, 2022. Daily COVID-19 Dashboard - Ottawa Public Health [WWW Document]. URL https://www.ottawapublichealth.ca/en/reports-research-and-statistics/daily-covid19-dashboard.aspx (accessed 5.26.22).

Paredes, M.I., Lunn, S.M., Famulare, M., Frisbie, L.A., Painter, I., Burstein, R., Roychoudhury, P., Xie, H., Bakhash, S.A.M., Perez, R., Lukes, M., Ellis, S., Sathees, S., Mathias, P.C., Greninger, A., Starita, L.M., Frazar, C.D., Ryke, E., Zhong, W., Gamboa, L., Threlkeld, M., Lee, J., McDermot, E., Truong, M., Nickerson, D.A., Bates, D.L., Hartman, M.E., Haugen, E., Nguyen, T.N., Richards, J.D., Rodriguez, J.L., Stamatoyannopoulos, J.A., Thorland, E., Melly, G., Dykema, P.E., MacKellar, D.C., Gray, H.K., Singh, A., Peterson, J.M., Russell, D., Torres, L.M., Lindquist, S., Bedford, T., Allen, K.J., Oltean, H.N., 2021. Associations between SARS-CoV-2 variants and risk of COVID-19 hospitalization among confirmed cases in Washington State: a retrospective cohort study. medRxiv. https://doi.org/10.1101/2021.09.29.21264272

Peccia, J., Zulli, A., Brackney, D.E., Grubaugh, N.D., Kaplan, E.H., Casanovas-Massana, A., Ko, A.I., Malik, A.A., Wang, D., Wang, M., Warren, J.L., Weinberger, D.M., Arnold, W., Omer, S.B., 2020. Measurement of SARS-CoV-2 RNA in wastewater tracks community infection dynamics. Nat. Biotechnol. 38, 1164–1167. https://doi.org/10.1038/s41587-020-0684-z

Planas, D., Veyer, D., Baidaliuk, A., Staropoli, I., Guivel-Benhassine, F., Rajah, M.M., Planchais, C., Porrot, F., Robillard, N., Puech, J., Prot, M., Gallais, F., Gantner, P., Velay, A., Le Guen, J., Kassis-Chikhani, N., Edriss, D., Belec, L., Seve, A., Courtellemont, L., Péré, H., Hocqueloux, L., Fafi-Kremer, S., Prazuck, T., Mouquet, H., Bruel, T., Simon-Lorière, E., Rey, F.A., Schwartz, O., 2021. Reduced sensitivity of SARS-CoV-2 variant Delta to antibody neutralization. Nature 596, 276–280. https://doi.org/10.1038/s41586-021-03777-9

Prevost, B., Lucas, F.S., Ambert-Balay, K., Pothier, P., Moulin, L., Wurtzer, S., 2015. Deciphering the diversities of astroviruses and noroviruses in wastewater treatment plant effluents by a high-throughput sequencing method. Appl. Environ. Microbiol. 81, 7215–7222. https://doi.org/10.1128/AEM.02076-15

Public Health Ontario, 2022a. Ontario COVID-19 Data Tool | Public Health Ontario [WWW Document]. URL https://www.publichealthontario.ca/en/data-and-analysis/infectious-disease/covid-19-data-surveillance/covid-19-data-tool?tab=vaccine (accessed 5.27.22).

Public Health Ontario, 2022b. Enhanced Epidemiological Summary - Early Dynamics of Omicron in Ontario November 1 to December 23, 2021.

Public Health Ontario, 2022c. COVID-19 Omicron (B.1.1.529) Variant of Concern and Communicability … What We Know So Far 1–15.

Randazzo, W., Truchado, P., Cuevas-Ferrando, E., Simón, P., Allende, A., Sánchez, G., 2020. SARS-CoV-2 RNA in wastewater anticipated COVID-19 occurrence in a low prevalence area. Water Res. 181. https://doi.org/10.1016/j.watres.2020.115942

Rimoldi, S.G., Stefani, F., Gigantiello, A., Polesello, S., Comandatore, F., Mileto, D., Maresca, M., Longobardi, C., Mancon, A., Romeri, F., Pagani, C., Cappelli, F., Roscioli, C., Moja, L., Gismondo, M.R., Salerno, F., 2020. Presence and infectivity of SARS-CoV-2 virus in wastewaters and rivers. Sci. Total Environ. 744, 140911. https://doi.org/10.1016/J.SCITOTENV.2020.140911

Saguti, F., Magnil, E., Enache, L., Churqui, M.P., Johansson, A., Lumley, D., Davidsson, F., Dotevall, L., Mattsson, A., Trybala, E., Lagging, M., Lindh, M., Gisslén, M., Brezicka, T., Nyström, K., Norder, H., 2021. Surveillance of wastewater revealed peaks of SARS-CoV-2 preceding those of hospitalized patients with COVID-19. Water Res. 189, 116620. https://doi.org/10.1016/J.WATRES.2020.116620

Sah, P., Fitzpatrick, M.C., Zimmer, C.F., Abdollahi, E., Juden-Kelly, L., Moghadas, S.M., Singer, B.H., Galvani, A.P., 2021. Asymptomatic SARS-CoV-2 infection: A systematic review and meta-analysis. Proc. Natl. Acad. Sci. U. S. A. 118. https://doi.org/10.1073/PNAS.2109229118/-/DCSUPPLEMENTAL

Santiso-Bellón, C., Randazzo, W., Pérez-Cataluña, A., Vila-Vicent, S., Gozalbo-Rovira, R., Muñoz, C., Buesa, J., Sanchez, G., Díaz, J.R., 2020. Epidemiological Surveillance of Norovirus and Rotavirus in Sewage (2016–2017) in Valencia (Spain). Microorganisms 8, 458. https://doi.org/10.3390/MICROORGANISMS8030458

Sherchan, S.P., Shahin, S., Ward, L.M., Tandukar, S., Aw, T.G., Schmitz, B., Ahmed, W., Kitajima, M., 2020. First detection of SARS-CoV-2 RNA in wastewater in North America: A study in Louisiana, USA. Sci. Total Environ. 743, 140621. https://doi.org/10.1016/J.SCITOTENV.2020.140621

Smith-Jeffcoat, S.E., Pomeroy, M.A., Sleweon, S., Sami, S., Ricaldi, J.N., Gebru, Y., Walker, B., Brady, S., Christenberry, M., Bart, S., Vostok, J., Meyer, S., Seys, S., Markelz, A., Ditto, N., Newbern, V., Thomas, F.J., Thomas, D., Cabredo, E., Kellner, S., Brown, V.R., Tate, J.E., Kirking, H.L., 2022. Multistate Outbreak of SARS-CoV-2 B.1.1.529 (Omicron) Variant Infections Among Persons in a Social Network Attending a Convention — New York City, November 18–December 20, 2021. Morb. Mortal. Wkly. Rep. 71, 238–242. https://doi.org/10.15585/mmwr.mm7107a3

Tartof, S.Y., Slezak, J.M., Fischer, H., Hong, V., Ackerson, B.K., Ranasinghe, O.N., Frankland, T.B., Ogun, O.A., Zamparo, J.M., Gray, S., Valluri, S.R., Pan, K., Angulo, F.J., Jodar, L., McLaughlin, J.M., 2021. Effectiveness of mRNA BNT162b2 COVID-19 vaccine up to 6 months in a large integrated health system in the USA: a retrospective cohort study. Lancet 398, 1407–1416. https://doi.org/10.1016/S0140-6736(21)02183-8

Wang, Y., Zhang, L., Li, Q., Liang, Z., Li, T., Liu, S., Cui, Q., Nie, J., Wu, Q., Qu, X., Huang, W., 2022. The significant immune escape of pseudotyped SARS-CoV-2 variant Omicron. Emerg. Microbes Infect. 11, 1–5. https://doi.org/10.1080/22221751.2021.2017757/SUPPL_FILE/TEMI_A_2017757_SM9424.JPG

WHO, 2021. Classification of Omicron (B.1.1.529): SARS-CoV-2 Variant of Concern [WWW Document]. URL https://www.who.int/news/item/26-11-2021-classification-of-omicron-(b.1.1.529)-sars-cov-2-variant-of-concern (accessed 6.1.22).

Wolter, N., Jassat, W., DATCOV-Gen author group, Gottberg, A. von, Cohen, C., 2022. Clinical severity of Omicron sub-lineage BA.2 compared to BA.1 in South Africa. medRxiv 2022.02.17.22271030. https://doi.org/10.1101/2022.02.17.22271030

Wu, F., Xiao, A., Zhang, J., Moniz, K., Endo, N., Armas, F., Bushman, M., Chai, P.R., Duvallet, C., Erickson, T.B., Foppe, K., Ghaeli, N., Gu, X., Hanage, W.P., Huang, K.H., Lee, W.L., Matus, M., McElroy, K.A., Rhode, S.F., Wuertz, S., Thompson, J., Alm, E.J., 2021. Wastewater Surveillance of SARS-CoV-2 across 40 U.S. states. medRxiv 1–13. https://doi.org/10.1101/2021.03.10.21253235

Wurtzer, S., Marechal, V., Mouchel, J., Maday, Y., Teyssou, R., Richard, E., Almayrac, J., Moulin, L., 2020. Evaluation of lockdown impact on SARS-CoV-2 dynamics through viral genome quantification in Paris wastewaters. medRxiv 2020.04.12.20062679. https://doi.org/10.1101/2020.04.12.20062679

Xiao, A., Wu, F., Bushman, M., Zhang, J., Imakaev, M., Chai, P.R., Duvallet, C., Endo, N., Erickson, T.B., Armas, F., Arnold, B., Chen, H., Chandra, F., Ghaeli, N., Gu, X., Hanage, W.P., Lee, W.L., Matus, M., McElroy, K.A., Moniz, K., Rhode, S.F., Thompson, J., Alm, E.J., 2021. Metrics to relate COVID-19 wastewater data to clinical testing dynamics. medRxiv 6, 2021.06.10.21258580. https://doi.org/10.1101/2021.06.10.21258580

Zhang, Y., Cen, M., Hu, M., Du, L., Hu, W., Kim, J.J., Dai, N., 2021. Prevalence and Persistent Shedding of Fecal SARS-CoV-2 RNA in Patients With COVID-19 Infection: A Systematic Review and Meta-analysis. Clin. Transl. Gastroenterol. 12, e00343. https://doi.org/10.14309/CTG.0000000000000343

