## Supplemental Material for "Understanding the dynamic relation between wastewater SARS-CoV-2 signal and clinical metrics throughout the pandemic"

**Dr. Robert Delatolla**

Work

### Supplemental Material

**Table S1:** List of PCR primer and probe sets utilized in this study

| Primer/probe & supplier | Sequence | Reference |
| --- | --- | --- |
| 2019-nCov_N1 forward primer (IDT) | GAC CCC AAA ATC AGC GAA AT | (CDC, 2020) |
| 2019-nCoV_N1 reverse primer (IDT) | TCT GGT TAC TGC CAG TTG AAT CTG | (CDC, 2020) |
| 2019-nCoV_N1 probe (IDT) | 6-FAM-ACC CCG CAT/ZEN/ TAC GTT TGG TGG ACC-IOWA BLACK FQ | (CDC, 2020) |
| 2019-nCoV_N2 forward primer (IDT) | TTA CAA ACA TTG GCC GCA AA | (CDC, 2020) |
| 2019-nCoV_N2 reverse primer (IDT) | GCG CGA CAT TCC GAA GAA | (CDC, 2020) |
| 2019-nCoV_N2 probe (IDT) | 6-FAM-ACA ATT TGC/ZEN/CCC CAG CGC TTC AG-IOWA BLACK FQ | (CDC, 2020) |
| PMMoV forward primer (ABI) | GAG TGG TTT GAC CTT AAC GTT GA | (Lee *et al*., 2018) |
| PMMoV reverse primer (ABI) | TTG TCG GTT GCA ATG CAA GT | (Lee *et al*., 2018) |
| PMMoV probe (ABI) | 6-FAM-CCT ACC GAA GCA AAT G-MGB | (Lee *et al*., 2018) |

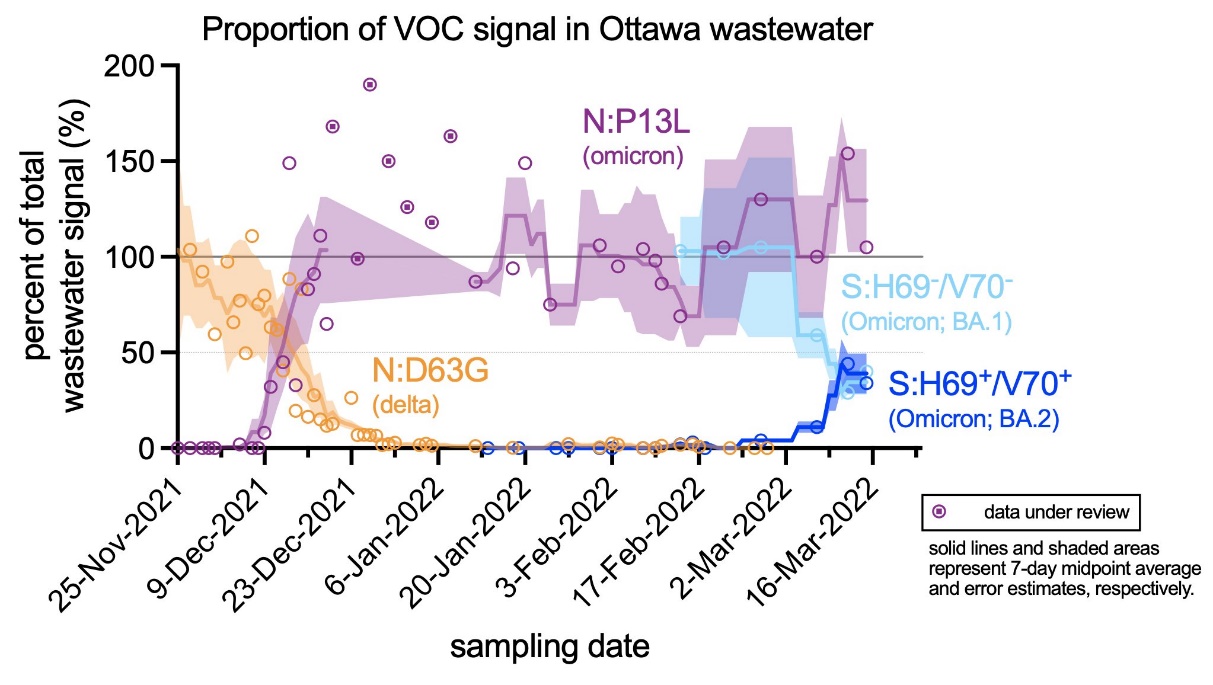

**Figure S1:** Proportions of B.1.617.2 (Delta), B.1.1.529.1 (Omicron BA.1), and B.1.1.529.2 (Omicron BA.2) VOC found in Ottawa wastewater between Nov. 25^th^, 2021 to Mar. 16^th^, 2022

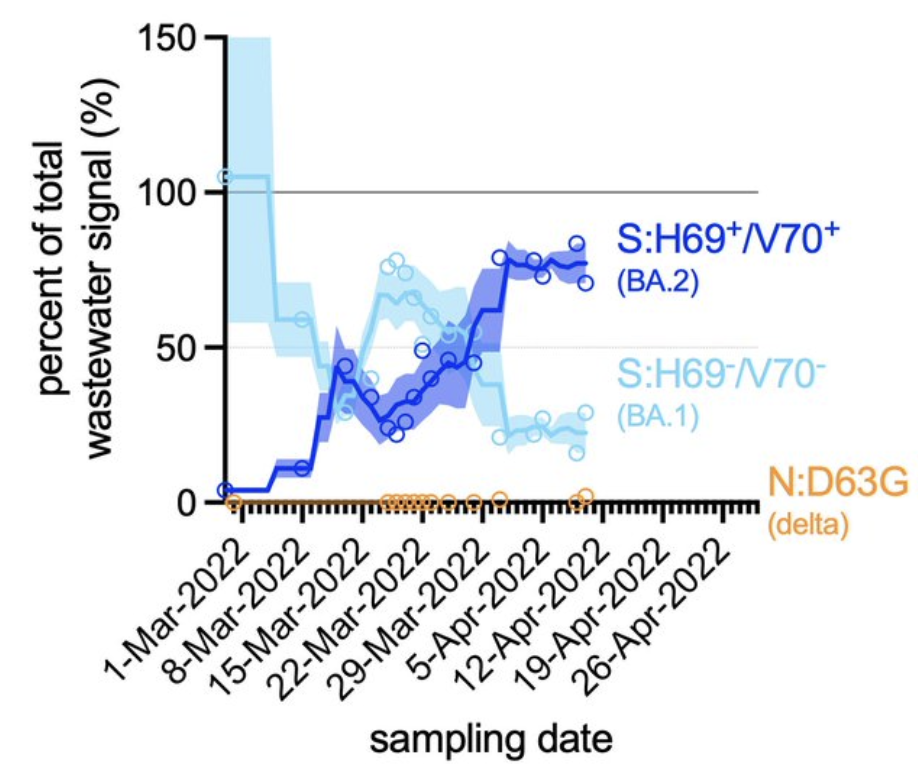

**Figure S2:** Proportions of B.1.617.2 (Delta), B.1.1.529.1 (Omicron BA.1), and B.1.1.529.2 (Omicron BA.2) VOC found in Ottawa wastewater between Mar. 1^st^, 2022 to Apr. 10^th^, 2022
